# Phenotypic Characteristics Associated with Preterm Births in the Indian Population

**DOI:** 10.1101/2025.05.18.25327852

**Authors:** Veerendra P. Gadekar, Aditi Sadhu, Shambho Basu Thakur, Ayushi, Rahul Jain, Dharmendra Sharma, Umesh Mehta, Alka Singh, GARBH-Ini Study Group, Nitya Wadhwa, Shinjini Bhatnagar, Siddarth Ramji, Ramachandran Thiruvengadam, Himanshu Sinha

## Abstract

**Background:** Preterm birth (PTB), defined as the birth of infants before 37 weeks of gestation, poses significant health risks, including increased mortality and long-term disabilities. India stands as a major contributor to global PTB-related mortality, primarily due to its substantial prevalence. The complex temporal relationship among multiple etiological factors makes predicting PTB occurrence challenging. This is further complicated by the diverse PTB phenotypic categories, namely maternal, foetal, placental, parturition, and pathway-to-delivery features affecting birth outcomes.

**Methods:** Our study focused on PTB in the Indian population using a comprehensive dataset from the GARBH-Ini cohort. We analysed 21 variables across maternal, foetal, placental, parturition-related, and other phenotypic categories in the context of Phenotype I (PTB/Non-PTB) and Phenotype II, which classifies PTB based on the pathway to delivery as either Spontaneous Onset of Labour (SOL) or Caregiver-Initiated (CGI). We assessed the prevalence of these phenotypic variables using the complete dataset and applied hierarchical clustering to identify key PTB-associated phenotypic patterns and their impact on neonatal outcomes, including NICU admissions and early neonatal mortality. Additionally, we used one-year follow-up data to examine infant-related phenotypic variables and their distribution across PTB and non-PTB cases.

**Results:** We identified key phenotypic variables across maternal, foetal, placental, and parturition categories that significantly influenced PTB outcomes. In SOL cases, foetal (perinatal sepsis, polyhydramnios) and parturition-related (PROM, short cervix, peripartum bleeding) features were significantly associated. In contrast, CGI cases were predominantly associated with maternal complications like pre-eclampsia. Placental abnormalities, such as placenta previa, in combination with PROM and short cervix, were notably prevalent in PTB cases, impacting neonatal mortality, NICU admissions, and infant health.

**Conclusions:** The study underscores the complexity of PTB as a time-based, multifaceted syndrome driven by diverse etiological factors. India’s prominence in global PTB-related mortality necessitates tailored preventive strategies. Our findings emphasise the significance of monitoring specific phenotype class and their associated features, facilitating the development of targeted interventions to reduce PTB occurrences in the Indian population. These relationships among features could assist in prioritising cases for personalised care and improve understanding of how PTB phenotypes affect infant health.

## INTRODUCTION

It is estimated that around 15 million babies are born preterm every year, of whom a million children die each year due to complications of preterm [1]. Many survivors face the risk of a lifetime of disability, including learning disabilities and visual and hearing problems. Based on the countries with reliable trend data, PTB rates have not declined despite socioeconomic development [2]. It is estimated that over 60% of PTB worldwide comes from Sub-Saharan Africa and South Asia [3], and India alone is the most significant contributor to the global burden [4, 5]. While the definition of PTB is on a time scale, defined as live birth <37 weeks of gestation, the characteristics of this adverse event are heterogeneous. The diversity in etiologic factors and events that occur during the perinatal period, collectively put together as a ‘phenotype,’ contributes to the heterogeneity in PTB and influences neonatal and infantile outcomes of a PTB [6, 7].

In this context, a framework has been proposed to classify PTB phenotypes based on maternal, foetal, and placental conditions, signs of parturition, and the PTB pathway to delivery [8]. A cross-sectional study of the INTERGROWTH-21st utilised this framework to classify and identify PTB phenotypes [9]. They identified 12 PTB phenotypes associated with different patterns of neonatal outcomes. Such types and distribution of PTB phenotypes might vary from population to population.

In this study, we have identified PTB phenotypes using maternal, foetal, placental conditions, and pathway to delivery, as these groups may have distinct underlying causes, risk factors, and clinical trajectories.[10] Given that a pregnancy is influenced by a multitude of factors causing significant variations between pregnancies, we adopted a hierarchical clustering approach, which allowed us to compare smaller groups of similar pregnancies, creating an opportunity to identify key phenotypic categories and their combinations critical to PTB outcomes.

## METHODS

### Study design and data collection

For the present study, data available in the GARBH-Ini cohort was used. This hospital-based pregnancy cohort commenced in May 2015 at Gurugram Civil Hospital, Gurugram, Haryana, India. Women were enrolled before 20 weeks of pregnancy and followed till delivery and once post-partum. For each participant, a wide array of data was collected, including maternal anthropometric measurements, clinical and obstetric histories, socioeconomic information, various biospecimens, and maternal and foetal biometry ultrasound assessments. The detailed methodologies of the GARBH-Ini study have been documented in a prior publication [11].

The institutional ethics committees of Gurugram Civil Hospital, Safdarjung Hospital (New Delhi), the Translational Health Science and Technology Institute, and the Indian Institute of Technology Madras approved the study.

### Dataset preparation and phenotypic variable definition

We used 21 variables (maternal, foetal, placental features, signs of parturition, and others) from the GARBH-Ini cohort for analysis (Table S2). These variables were selected based on their relevance to the phenotypic classification framework proposed by Villar et al.[8], and were critical for analysing their relationship with PTB outcomes. We obtained data from 10494 participants enrolled in the cohort. For this study, we only selected data from participants who had delivered at the Gurugram Civil Hospital, with available details of delivery and pregnancy outcome (N = 5232). We excluded stillbirths and those who had delivered at < 28 weeks gestational age (numbers were too small to make inferences with certainty, Figure 1A).

**Figure 1.**
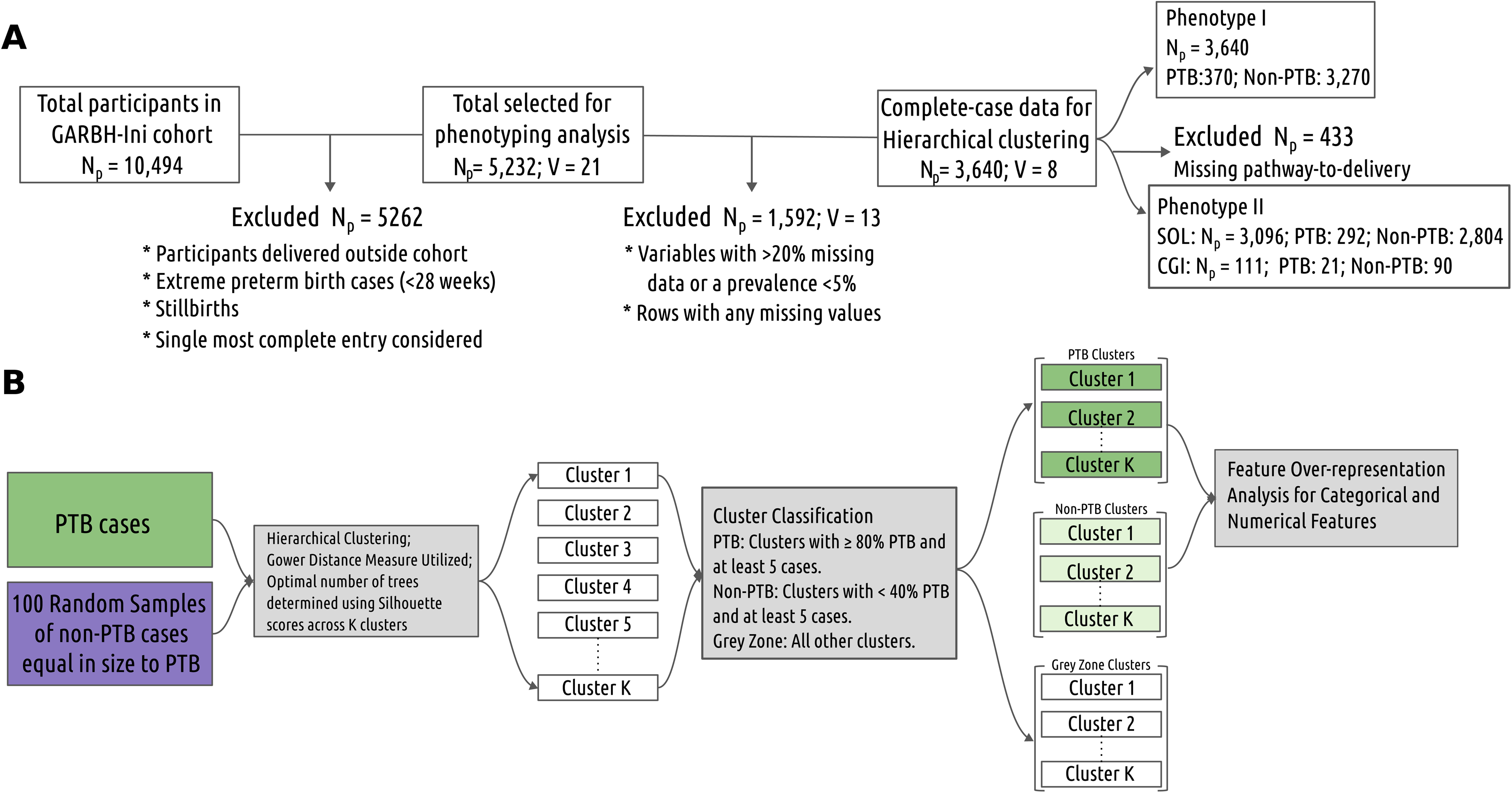
Overview of data selection and clustering workflow used for PTB phenotyping. **(A)** A stepwise data selection process was used to construct the final dataset (N = 3640) for clustering analysis. This included the application of inclusion/exclusion criteria to phenotypic variables from the GARBH-Ini cohort to obtain a complete-case subset for clustering. **(B)** Clustering workflow applied to the selected dataset, including preprocessing, distance metric selection, hierarchical clustering, and cluster labelling. This workflow was used to identify clinically meaningful subgroups based on phenotypic patterns associated with PTB.

### Exploratory data analysis

We compared the proportion of PTB and non-PTB cases for each variable using a one-tailed Fisher’s exact test, with the alternative hypothesis set to ‘greater.’ P-values were adjusted for multiple comparisons using the Bonferroni method. A variable was considered significantly more prevalent in PTB cases if the adjusted P-value was < 0.05. In addition, we assessed pairwise associations between variables by calculating the proportion of total cases in the PTB and non-PTB groups where one variable was recorded as ‘yes,’ given that another variable was also recorded as ‘yes.’

### Hierarchical clustering

For the clustering analysis, based on our exploratory assessment (Figures S1, S2), we selected variables with less than 20% missing data and a prevalence of at least 5%. Variables with lower prevalence were excluded, as they typically contribute little to meaningful cluster differentiation. Based on these criteria, eight variables were chosen (five maternal and one each from foetal, placental, and other categories, Table S4). Additionally, we included two continuous numerical variables, cervix length and amniotic fluid index, as their continuous nature provides finer granularity and can enhance the clustering process by capturing subtle differences between data points. Rows with missing values in the selected variables were removed, resulting in a complete-case dataset comprising 3640 participants. This included 370 (10.16%) PTB cases and 3270 (89.83%) non-PTB cases and was used for PTB phenotype analysis. It also contained 3096 SOL cases (85.05%) and 111 CGI cases (3.04%), which were subsequently used to analyse PTB phenotypes based on delivery pathway. PTB was recorded in 292 (9.43%) of the SOL cases and 21 (18.91%) of the CGI cases.

To identify distinct clusters of PTB and non-PTB cases, we performed agglomerative (bottom-up) hierarchical clustering using both binary and continuous variables. Given the imbalance between PTB and non-PTB cases, we conducted 100 random samplings, selecting an equal number of cases from each group in every iteration to enhance the robustness of our analysis (Figure 1B). Since our dataset contained binary and continuous numeric variables, we used the Gower distance metric [12], which normalises variables based on their respective ranges, ensuring comparability across different data types. This metric functions similarly to the Manhattan distance but incorporates range normalisation.

To evaluate the quality of clustering, we used silhouette scores [13] ranging from −1 to +1, with higher values indicating more distinct and well-defined clusters (Supplementary Information; Figure S5). By analysing the distribution of average silhouette scores across varying numbers of clusters (*K*) and their sizes, we identified *K* = 20 as the optimal solution, yielding an average silhouette score of ∼0.6 across all three datasets used for Phenotype I (also referred to as PTB Phenotype) and Phenotype II (corresponding to the SOL and CGI subset) analysis. The average median cluster sizes were 13.82 for the full dataset (N = 3640), 10.56 for the SOL subset (N = 3096), and 1.32 for the CGI subset (N = 111). Clustering was implemented in R using the distmix() function from the kmed package [14] (v0.4.2) and the hclust() function from the stats package (v4.1.2).

### Classification of the clusters

A cluster with at least 5 cases was labelled a PTB cluster if 80% or more were PTB cases. Similarly, a cluster with at least 5 cases was labelled a non-PTB cluster if less than 40% of its cases were PTB. Clusters that did not meet either of these conditions were considered “grey zone” (Figure 1) and excluded from further analysis. On average, in 100 random samples, the majority of clusters fell into this grey zone: about 14 out of 20 clusters in the full dataset, 16 out of 20 in the SOL dataset, and 18 out of 20 in the CGI dataset. This filtering step allowed us to retain optimally sized clusters for statistical testing and compare the prevalence of variables between PTB and non-PTB clusters.

### Assessing the prevalence of phenotypic variables in PTB clusters

After identifying PTB and non-PTB clusters and their corresponding enrolment IDs, we mapped all 21 variables (Table S2) across all clusters in 100 random samples. We first assessed the normality of the distribution of the proportion of cases marked ‘yes’ for each feature. Since the distributions were non-normal, we applied a one-sided Wilcoxon test to determine whether the proportion of cases marked ‘yes’ was higher in PTB than in non-PTB cases. The null hypothesis (HLJ) assumed no difference between the groups, meaning the median proportions were equal. To account for multiple comparisons, we adjusted P-values using the Bonferroni method, implemented in R through a custom in-house script. Effect size was quantified using Fold Change (FC), calculated as the ratio of the proportion in the PTB group (foreground) to that in the non-PTB group (background). Features were prioritised based on FC and an adjusted P-value threshold (P_adj_ < 0.05), ranking them according to their relevance to PTB outcomes. A feature was considered overrepresented in PTB clusters if it showed P_adj_ < 0.05. Additionally, we extended this analysis to assess pairwise feature combinations.

### Analysing the newborn features with variables of PTB

We analysed the relationship between preterm infant features and the selected variables, independent of the clustering-based analysis. Through pairwise comparisons, infant features were examined in relation to maternal, foetal, placental, and parturition characteristics. For each pair, we assessed the number of PTB cases where both the infant feature and the phenotypic variable were marked as ‘yes.’ These cases formed the foreground group, representing the co-occurrence of the two features. The background group comprised the remaining PTB cases where either one or neither of the features was marked as ‘yes.’ A one-sided Fisher’s exact test (alternative = “greater”) was used to evaluate whether co-occurrence was significantly enriched compared to the background. FC was calculated as the ratio of the proportion of cases in the foreground to that in the background. Bonferroni correction was applied to account for multiple testing within each dataset. This analysis was performed separately for the full dataset (N = 5129), the SOL subset (N = 4324), and the CGI subset (N = 172), enabling the identification of both consistent and subset-specific associations between infant outcomes and phenotypic features of PTB.

## RESULTS

The baseline characteristics of participants included and excluded from this study are provided in Table 1 and Table S1.

**Table 1:** Baseline characteristics of the participants included in the study.

| **Sociodemographic characteristics** | **Selected Participants (N = 5231) set median (IQR) or N (%) or mean ± SD** |
| --- | --- |
| Age (years) | 24 (21, 26) |
| **BMI at enrolment into the cohort** |  |
| Underweight | 24.90% |
| Normal | 58.47% |
| Overweight | 12.59% |
| Obese | 2.42% |
| Haemoglobin (g/dL) | 9.2 (8.6–10.4) |
| Weight (kgs) | 58 |
| **Socioeconomic status** |  |
| 0 | 0.26% |
| 1 | 11.01% |
| 2 | 26.34% |
| 3 | 38.48% |
| 4 | 0.82% |
| Undetermined | 23.07% |
| **Level of education** |  |
| Illiterate | 16.97% |
| Literate or primary school | 8.96% |
| Middle school | 16.34% |
| High school | 19.02% |
| Post-high school diploma | 21.46% |
| Graduate | 14.01% |
| Post-graduate | 3.21% |
| Unknown | 0.00% |
| **Occupation** |  |
| Unemployed | 91.35% |
| Unskilled worker | 3.80% |
| Semi-skilled worker | 1.62% |
| Skilled worker | 2.46% |
| Clerk, shop, farm owner | 0.30% |
| Semi-professional | 0.22% |
| Professional | 0.21% |
| Unknown | 0.00% |
| **History of any chronic illnesses** |  |
| Absent | 97.03% |
| Present | 2.98% |

Of the 5232 selected participants, 609 (11.63%) experienced PTB. Pathway to delivery information was available for 4656 participants (88.99%). Among them, 4390 (94.24%) had SOL, while the remaining 175 had CGI deliveries. Among those with SOL, 472 (10.75%) reported PTB, whereas 31 (1.77%) of the CGI group experienced PTB. Exploratory data analysis was conducted on all the selected variables (Table S2) to assess their distributions and the proportion of missing data (Figures S1, S2, S4).

We also identified features related to newborn conditions using follow-up information available for 5129 participants (98.03%). Of these, 579 (11.28%) were PTB. The infant condition-related features are presented in Table S3, and the results of the exploratory analysis are shown in Figure S3.

### Prevalence of characteristics in the PTB phenotype

Pre-eclampsia, short cervix, hypertension, perinatal sepsis, history of previous PTB among multiparous women, polyhydramnios, oligohydramnios, and PROM were significantly more prevalent in PTB than in the non-PTB group. The prevalence of all characteristics in the PTB phenotype is summarised in Table 2.

**Table 2:** Enrichment of phenotypic variables in PTB versus non-PTB cases in the complete dataset (PTB-Phenotype, N = 5232). The table presents the number and percentage of PTB and non-PTB cases for each phenotypic variable, grouped by clinical category. Statistical significance indicates overrepresentation of the variable in PTB compared to non-PTB cases, based on adjusted P-values corrected (Bonferroni method) for multiple testing. ** indicates P_adj_ < 0.01; * indicates P_adj_ < 0.05; variables without asterisks are not statistically significant.

| Class | Phenotypic Variables | # PTB (%) | # Non-PTB (%) | Adjusted P-value |
| --- | --- | --- | --- | --- |
| Maternal | **Short Cervix \*\*** | 66 (10.84%) | 158 (3.42%) | 1.84E-12 |
| Maternal | Chorioamnionitis | 6 (0.99%) | 37 (0.8%) | 1.00E+00 |
| Maternal | Bacterial vaginosis | 35 (5.75%) | 300 (6.49%) | 1.00E+00 |
| Maternal | Hepatitis | 9 (1.48%) | 41 (0.89%) | 1.00E+00 |
| Maternal | Respiratory infections | 50 (8.21%) | 377 (8.15%) | 1.00E+00 |
| Maternal | UTI | 197 (32.35%) | 1522 (32.92%) | 1.00E+00 |
| Maternal | Eclampsia ** | 58 (9.52%) | 239 (5.17%) | 4.98E-04 |
| Maternal | Medical Disorders | 423 (69.46%) | 3096 (66.97%) | 1.00E+00 |
| Maternal | Anaemia * | 277 (45.48%) | 1932 (41.79%) | 6.85E-01 |
| Maternal | Hyperthyroidism | 13 (2.13%) | 35 (0.76%) | 3.82E-02 |
| Maternal | Hypothyroidism | 31 (5.09%) | 148 (3.2%) | 2.12E-01 |
| Maternal | Parity | 380 (62.4%) | 2804 (60.65%) | 1.00E+00 |
| Maternal | **Perinatal Sepsis \*\*** | 101 (16.58%) | 168 (3.63%) | 6.40E-29 |
| Maternal | **PTB History \*\*** | 73 (19.21%) | 199 (7.1%) | 1.75E-11 |
| Maternal | Rupture of Uterine | 2 (0.33%) | 6 (0.13%) | 1.00E+00 |
| Foetal | IUGR | 163 (26.77%) | 1819 (39.35%) | 1.00E+00 |
| Foetal | Foetal Anomaly | 4 (0.66%) | 10 (0.22%) | 2.81E-01 |
| Foetal | **Oligohydramnios \*\*** | 10 (1.64%) | 3 (0.06%) | 3.52E-07 |
| Foetal | **Polyhydramnios \*** | 13 (2.13%) | 35 (0.76%) | 1.02E-02 |
| Placental | Placenta Previa | 356 (58.46%) | 3057 (66.13%) | 1.00E+00 |
| Parturition | **PROM \*\*** | 132 (21.67%) | 615 (13.3%) | 8.76E-08 |
| Other | Biomass Fuel | 65 (10.67%) | 395 (8.54%) | 5.03E-02 |

To assess the association of delivery pathway on PTB, a stratified analysis by SOL and CGI deliveries was carried out. Results highlighted contrasting phenotypic profiles across the two subtypes (Table 3). In the SOL subset, the prevalence of perinatal sepsis, short cervix, previous history of PTB, polyhydramnios, oligohydramnios, and PROM was significantly higher in the PTB group. In contrast, in the CGI subset, only pre-eclampsia was significantly higher in the PTB group.

**Table 3:** Enrichment of phenotypic variables in PTB cases stratified by pathway to delivery. A total of 4390 participants had SOL, and 175 had CGI deliveries. The table shows the number and percentage of PTB and non-PTB cases for each phenotypic variable within the SOL and CGI subgroups. Statistical significance reflects overrepresentation of the variable in PTB compared to non-PTB cases within each subgroup, based on P-values adjusted for multiple testing (Bonferroni method). ** indicates P_adj_ < 0.01; * indicates P_adj_ < 0.05; variables without asterisks are not statistically significant.

| Class | Phenotypic Variables | SOL |  |  | CGI |  |  |
| --- | --- | --- | --- | --- | --- | --- | --- |
|  |  | # PTB (%) | # Non-PTB (%) | Adjusted P-value | # PTB (%) | # Non-PTB (%) | Adjusted P-value |
| Maternal | Short Cervix | **61 (12.92%)**<br>** | 141 (3.6%) | 8.84E-14 | 1 (3.23%) | 5 (3.47%) | 1.00E+00 |
| Maternal | Chorioamnionitis | 5 (1.06%) | 36 (0.92%) | 1.00E+00 | 1 (3.23%) | 0 (0%) | 1.00E+00 |
| Maternal | Bacterial vaginosis | 23 (4.87%) | 267 (6.81%) | 1.00E+00 | 4 (12.9%) | 5 (3.47%) | 8.04E-01 |
| Maternal | Hepatitis | 9 (1.91%) | 36 (0.92%) | 7.01E-01 | 0 (0%) | 3 (2.08%) | 1.00E+00 |
| Maternal | Respiratory infections | 38 (8.05%) | 324 (8.27%) | 1.00E+00 | 3 (9.68%) | 11 (7.64%) | 1.00E+00 |
| Maternal | UTI | 159 (33.69%) | 1301 (33.21%) | 1.00E+00 | 11 (35.48%) | 49 (34.03%) | 1.00E+00 |
| Maternal | Pre-eclampsia | 27 (5.72%) | 174 (4.44%) | 1.00E+00 | **13 (41.94%)**<br>* | 21 (14.58%) | 1.74E-02 |
| Maternal | Anaemia | 227 (48.09%) | 1658 (42.32%) | 1.45E-01 | 6 (19.35%) | 47 (32.64%) | 1.00E+00 |
| Maternal | Hypertension | 5 (1.06%) | 23 (0.59%) | 1.00E+00 | 2 (6.45%) | 5 (3.47%) | 1.00E+00 |
| Maternal | Hypothyroidism | 14 (2.97%) | 111 (2.83%) | 1.00E+00 | 3 (9.68%) | 3 (2.08%) | 1.00E+00 |
| Maternal | Parity | 291<br>(61.65%) | 2288<br>(58.4%) | 1.00E+00 | 12 (38.71%) | 77 (53.47%) | 1.00E+00 |
| Maternal | Perinatal Sepsis | **80 (16.95%)**<br>** | 143 (3.65%) | 8.72E-24 | 5 (16.13%) | 10 (6.94%) | 1.00E+00 |
| Maternal | History of<br>previous PTB | **53 (18.21%)**<br>** | 153 (6.69%) | 1.52E-08 | 5 (41.67%) | 6 (7.79%) | 8.51E-02 |
| Maternal | Rupture of<br>Uterine | 1 (0.21%) | 3 (0.08%) | 1.00E+00 | 0 (0%) | 0 (0%) | 1.00E+00 |
| Foetal | IUGR | 122<br>(25.85%) | 1577<br>(40.25%) | 1.00E+00 | 13 (41.94%) | 64 (44.44%) | 1.00E+00 |
| Foetal | Foetal Anomaly | 4 (0.85%) | 6 (0.15%) | 6.53E-02 | 0 (0%) | 0 (0%) | 1.00E+00 |
| Foetal | Oligohydramnios | **8 (1.69%)** ** | 2 (0.05%) | 2.50E-06 | 1 (3.23%) | 1 (0.69%) | 1.00E+00 |
| Foetal | Polyhydramnios | **11 (2.33%)**<br>** | 25 (0.64%) | 3.88E-03 | 0 (0%) | 2 (1.39%) | 1.00E+00 |
| Placental | Placenta Previa | 287<br>(60.81%) | 2632<br>(67.18%) | 9.97E-01 | 15 (48.39%) | 81 (56.25%) | 8.41E-01 |
| Parturition | PROM | **107**<br>**(22.67%)** ** | 529 (13.5%) | 2.85E-07 | 11 (35.48%) | 38 (26.39%) | 2.09E-01 |
| Other | Biomass Fuel | 50 (10.59%) | 333 (8.5%) | 7.79E-02 | 4 (12.9%) | 15 (10.42%) | 4.44E-01 |

### Cluster-specific enrichment of key phenotypic variables in the subtypes of PTB

Hierarchical clustering-based enrichment analysis identified distinct phenotypic determinants of PTB (Figure S6, Table S10) and for the phenotypes stratified by delivery pathway (Figures 2A, 2B, Table S10).

**Figure 2:**
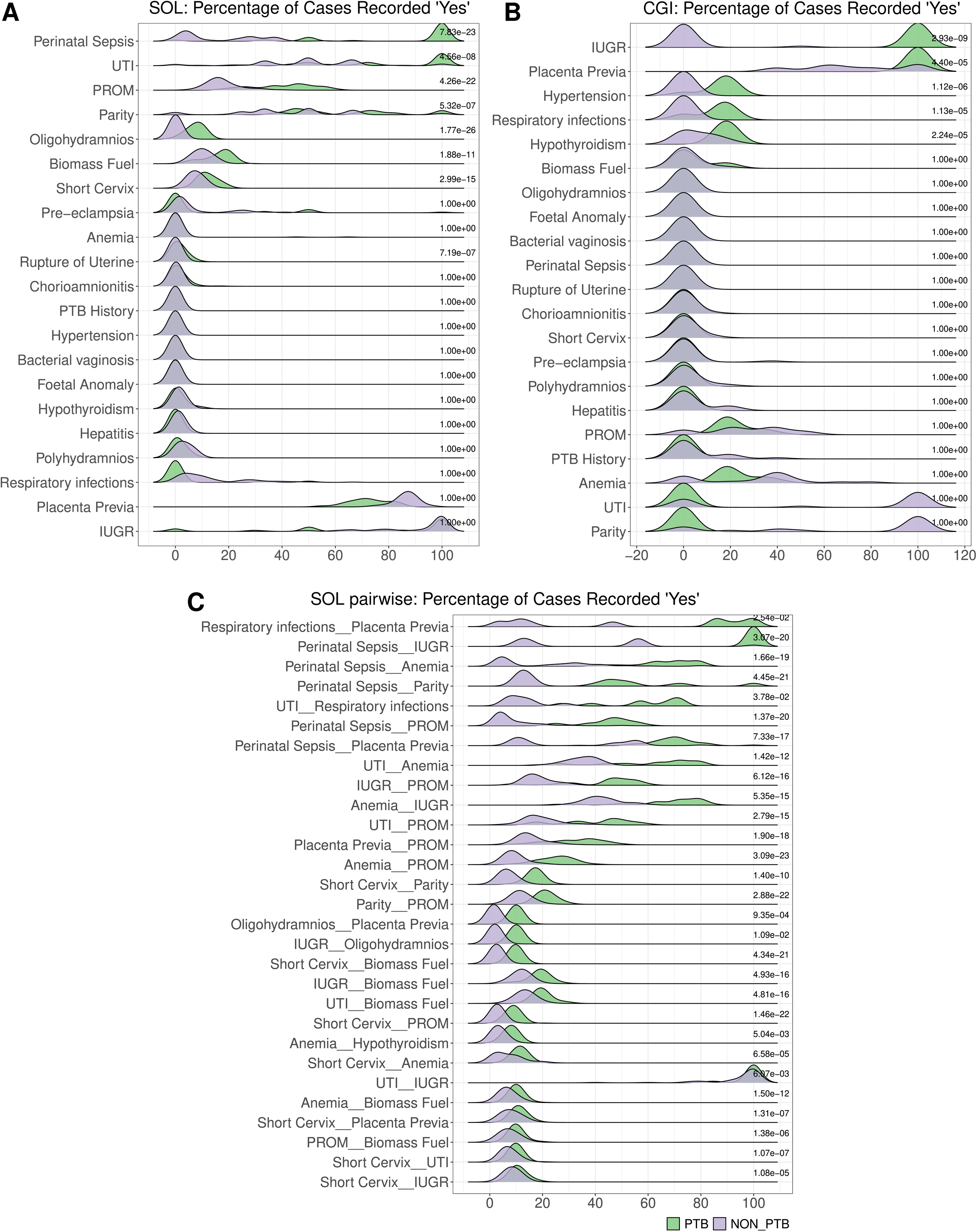
Distribution of the percentage of ‘Yes’ responses for phenotypic variables in PTB and non-PTB clusters using the SOL and CGI subsets. The figure shows the distribution of ‘Yes’ responses for the 21 phenotypic variables across clusters derived from two subsets of the dataset used in SOL and CGI analysis: **(A)** Spontaneous onset of labour (SOL; N = 3096) and **(B)** Clinician-initiated delivery (CGI; N = 111). **(C)** Distribution of significantly overrepresented co-occurring phenotypic variable pairs in PTB-dominant clusters within the SOL subset, selected for co-occurrence analysis based on variables identified as significant in panels A and B. Green denotes PTB-dominant clusters, and purple denotes non-PTB-dominant clusters. The distributions highlight differences in the prevalence of clinical phenotypic variables between PTB and non-PTB groups within each subset. Adjusted P-values for multiple comparisons are shown alongside each distribution to indicate the significance of observed differences.

In the PTB phenotype, the clusters exhibited significant enrichment of maternal, foetal, and parturition-related complications. Perinatal sepsis emerged as the most enriched variable, observed in 82.18% of PTB clusters versus 12.68% in non-PTB clusters (p < 0.001). Pre-eclampsia showed a pronounced increase in PTB clusters (17.47% versus 1.67%, p < 1e-17), along with other maternal features such as hypothyroidism (3.63% versus 1.92%, p < 0.001). Rare complications like uterine rupture (2.92%) and oligohydramnios (1.64% versus 0.06%, p < 0.001) were almost exclusively observed in PTB clusters, underscoring their clinical significance despite low prevalence. We also observed that PROM (21.67% versus 13.3%, p < 0.01) and short cervix (10.84% versus 3.42%, p < 0.01) were significantly enriched in PTB clusters, while placental abnormalities like placenta previa were less common in PTB cases.

In the SOL-PTB phenotype, perinatal sepsis remained the most prominent feature, enriched in 84.39% of PTB clusters compared to 16.44% in non-PTB clusters (p < 1e-22). Cervical insufficiency emerged as a key driver, with the short cervix significantly overrepresented in PTB clusters (12.25% versus 7.56%, p < 0.001), alongside PROM (40.76% versus 17.77%, p<0.001). Amniotic fluid abnormalities, such as oligohydramnios, showed substantial enrichment (7.53% versus 0.11%, p < 0.001), while maternal infections, like UTIs, were more frequent in PTB cases (76.39% versus 50.94%, p < 0.001).

In the CGI-PTB phenotype, the enrichment profile revealed hypertension (15.56% versus 0%, p < 0.001), placenta previa (100% versus 73.1%, p < 0.01), IUGR (100% versus 2.38%, p < 0.001), hypothyroidism (18.25% versus 6.27%, p < 0.001) and respiratory infections (13.49% versus 0%, p < 0.001) as the prominent factors for PTB.

### Pair-wise associations

An analysis of pairwise associations between phenotypic variables in PTB and non-PTB cases (Table S6) and in the SOL and CGI subsets (Tables S7, S8) was carried out to identify combinations of clinical features that co-occur more frequently in PTB. For the PTB phenotype, several pairs of characteristics showed more than twofold higher prevalence compared to non-PTB cases. For example, anaemia co-occurred with a short cervix in 10.11% of PTB versus 3.31% of non-PTB cases; with perinatal sepsis in 14.8% versus 3.47%, and with PROM in 23.47% versus 15.17%. Previous history of PTB occurred more frequently with anaemia (11.55% versus 4.40%), pre*-* eclampsia (15.52% versus 6.28%), and perinatal sepsis (13.79% versus 3.77%) in PTB than in non-PTB groups. Other important pairs seen in the PTB group include hypertension with PROM (30.77% versus 11.43%) and hypertension with short cervix (7.69% versus 2.03%); and hypothyroidism with short cervix (12.9% versus 2.03%) and hypothyroidism with PROM (22.58% versus 14.19%). In addition, IUGR co-occurred with short cervix in 10.43% of PTB compared to 3.42% of non-PTB cases.

In the SOL subset, several of these trends were preserved or further amplified. Polyhydramnios with a short cervix was present in 63.64% of PTB cases versus just 8.00% in non-PTB cases. Likewise, perinatal sepsis and PROM co-occurred in 80.00% of PTB versus 10.00% of non-PTB. Other notable pairings included PROM with a short cervix (17.76% versus 2.46%) and placenta previa with a short cervix (12.20% versus 3.00%). The history of previous PTB also remained a frequently enriched factor when combined with maternal infections and delivery complications. These patterns suggested that a short cervix, infections, and abnormal amniotic fluid levels might play a role in triggering spontaneous PTB.

Although the CGI subset was limited by a smaller sample size (only 31 PTB cases), a few high-prevalence pairings were observed. Maternal complications dominated this group. Pre-eclampsia co-occurred with chorioamnionitis and placenta previa exclusively in PTB cases, and perinatal sepsis with PROM was present in 80.00% of PTB versus 10.00% of non-PTB. Previous history of PTB appeared in enriched pairings with pre-eclampsia and anaemia.

In the pairwise analysis performed among the SOL subset, PTB clusters were notably enriched for infection-related combinations (Figure 2C, Table S11). All PTB cases in this group showed the presence of perinatal sepsis with IUGR. Other significantly over-represented combinations included respiratory infections with placenta previa, perinatal sepsis with anaemia, short cervix with PROM, UTI with PROM, and IUGR with PROM. Additionally, a short cervix combined with biomass fuel exposure was more frequently observed in PTB clusters. In contrast, the CGI subset did not show any statistically significant associations after P-value adjustment, likely due to smaller sample sizes or weaker correlations.

### Neonatal Outcomes

The analysis revealed significantly higher NICU admission and early neonatal death (END) rates in PTB cases compared to non-PTB cases. The variables with the highest NICU admission among PTB were perinatal sepsis (100%), hypertension (76.92%), and preeclampsia (62.07%), with the least being for anaemia (25.63%) and placenta previa (28.37%, Table S5).

### Association of PTB Phenotypes with Infant Clinical Outcomes

Using term deliveries as the reference group, we evaluated the association between infant clinical outcomes and PTB phenotypes (Table S9). Four conditions were significantly more prevalent in the PTB phenotype after multiple testing correction (P_adj_ < 0.01): respiratory distress, septic shock linked to perinatal sepsis, recurrent illness with anaemia, and CNS disorders associated with hypothyroidism. In the SOL-PTB phenotype, CNS disorders also showed a strong association with hypothyroidism. No significant associations were observed in the CGI-PTB phenotype, likely due to limited sample size. Overall, these findings highlight perinatal sepsis in the PTB phenotype as a major risk factor for respiratory and septic complications in infants, while hypothyroidism consistently co-occurred with infant CNS disorders in both PTB and SOL-PTB phenotypes.

## DISCUSSION

Our study demonstrates the heterogeneity of PTB phenotype (Figure 3) and identifies the most associated characteristics of this phenotype. PTB frequently presented with multiple co-occurring conditions, such as maternal conditions (e.g., pre-eclampsia, short cervix, perinatal sepsis, History of previous PTB, foetal factors (e.g., polyhydramnios, oligohydramnios), and parturition-related complications (e.g., PROM). CGI cases were primarily associated with maternal hypertensive disorders like pre-eclampsia, while SOL cases were more linked to perinatal sepsis and PROM.

**Figure 3:**
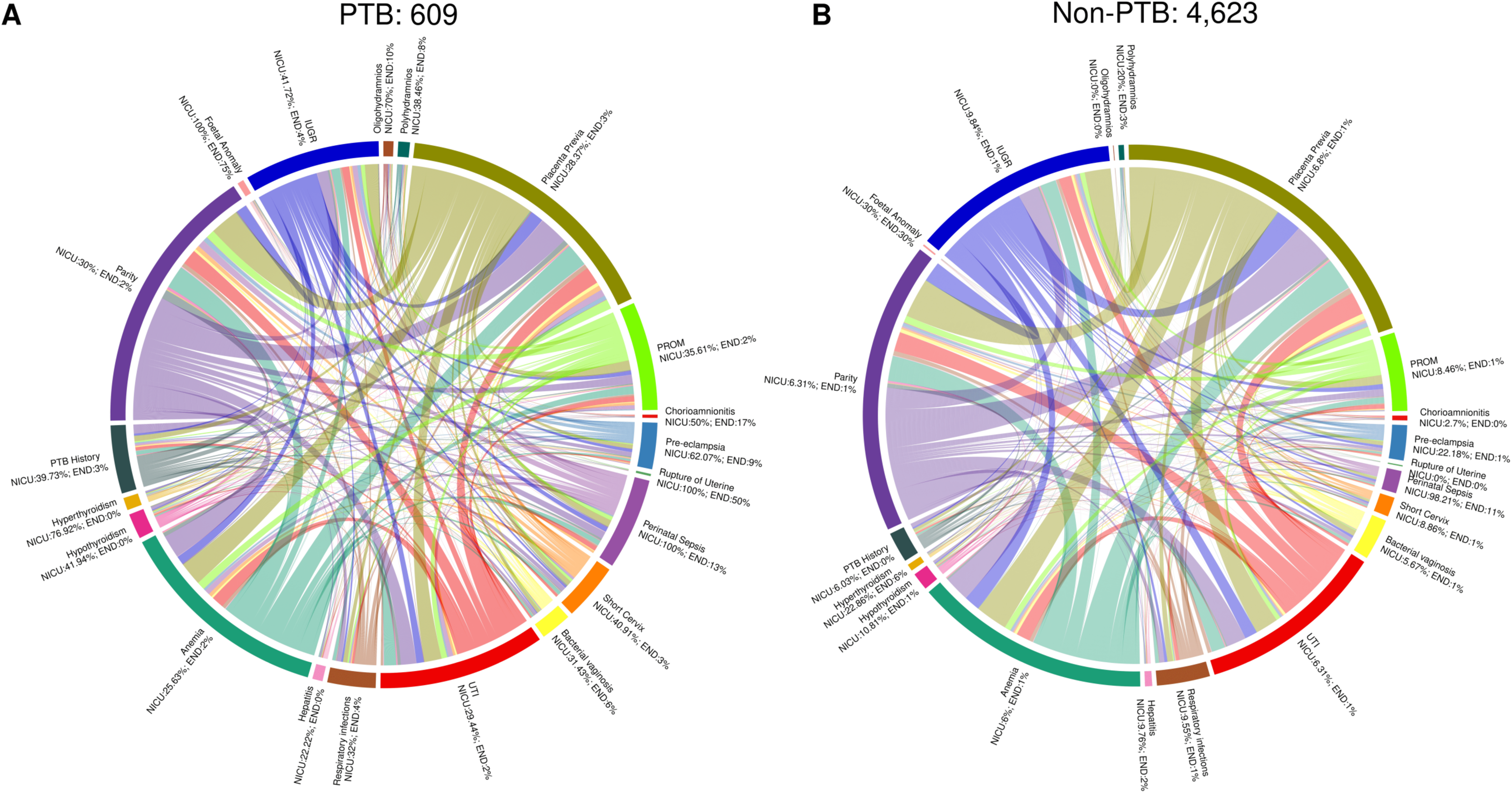
The figure illustrates the relationships and shared occurrences of various phenotypic variables among **(A)** PTB (left) and **(B)** non-PTB (right) cases. Each arc represents a distinct phenotypic variable, with connecting lines indicating co-occurrences with other variables. The thickness of the connecting lines and the proportional arc lengths reflect the frequency of co-occurrence across the dataset.

Many of these features that we demonstrate have been previously associated with PTB. For example, PROM is a well-known factor associated with PTB, contributing to about one-third of all preterm deliveries globally [15, 16], and it has also been associated with the development of intrauterine infections [17], which can lead to serious complications such as perinatal sepsis. In a prospective cohort study conducted in China, gestational hypertension and pre-eclampsia were found to be associated with a higher risk of PTB [18]. Women with a cervical length shorter than 25 mm (10th percentile) at 24 weeks faced a six-fold increased risk of spontaneous PTB before 35 weeks compared to those with lengths above 40 mm (75th percentile) [19]. Our findings underscore a complex interplay of phenotypic features strongly associated with PTB and the differences in distribution of neonatal outcomes in PTB with different phenotypic features.

The clustering-based enrichment analysis provided deeper insights into PTB by identifying distinct phenotypic clusters with co-occurring risk factors. In the SOL dataset, PTB-enriched clusters were primarily linked to maternal and parturition-related conditions such as perinatal sepsis, short cervix, and PROM. In contrast, the CGI dataset highlighted maternal complications, particularly IUGR and placenta previa, as dominant factors. These findings emphasise the diverse causes of PTB and the need to consider different risk factors for spontaneous and caregiver-induced cases.

Our results align with previous studies, which identified similar phenotypic categories affecting PTB outcomes [8, 9], demonstrating the consistency of phenotypes across populations. A notable convergence is the co-occurrence of specific maternal and foetal conditions, particularly perinatal sepsis, IUGR, and parity in the context of multiple births.

This is the first study in LMIC settings, such as India, in which the phenotype of PTB has been evaluated beyond traditional classification, such as spontaneous and caregiver-initiated subtypes. This study gives the profile of preterm births that occur in LMIC settings. The clinical applications of the phenotyping exercise are twofold. This helps us to identify characteristics, maternal, foetal, and placental, commonly associated with PTB. These characteristics, if found causal, can be targets for interventions to prevent PTB. Secondly, the greater advantage of phenotyping PTB is that it enables the development of prediction models targeted towards the phenotypes that are highly prevalent. Further, it enables prioritisation of PTB phenotypes that are strongly associated with the adverse neonatal outcomes.

## LIMITATIONS

First, the diversity of the population captured in this study represented only a subset of India’s broader demographic and ethnic landscape. Additionally, a subset of data was used because participants were lost to follow-up or delivered at sites outside the cohort. However, the risk of systematic loss-to-follow-up bias was minimal, as the baseline characteristics of participants included and excluded from the study were highly similar. Certain phenotypic features in the dataset had relatively lower frequencies, which may have limited the statistical power to detect significant differences.

Additionally, NICU admissions, which were used as an outcome in this study, could result from several factors not solely related to the phenotypic features under investigation. Finally, the associated phenotypic features are associations useful to characterise PTB and not intended to be interpreted as causal relationships.

## CONCLUSION

Our study highlights the key maternal, foetal, and parturition features and their combinations, such as placenta previa, perinatal sepsis, PROM, polyhydramnios, and medical disorders, which form a critical part of the PTB phenotype. Clinically, this detailed phenotyping can guide targeted interventions, enabling healthcare providers to offer appropriate prenatal care for high-risk pregnancies, ultimately improving maternal and neonatal outcomes. Expanding such studies with a more comprehensive dataset across diverse LMIC populations would further elucidate the PTB phenotype, providing a stronger foundation for the development of risk-stratification tools.

## Data Availability

The datasets used and analysed in the current study are available upon reasonable request from the corresponding author after approval of the DBT Steering Committee of GARBH-Ini.

## ACKNOWLEDGEMENTS

We thank all the participants of the GARBH-Ini study. We recognise the efforts of the research physicians, study nurses, clinical and laboratory technicians, field workers, the internal quality improvement team, and the project and data management team of the GARBH-Ini cohort. We thank members of the Centre for Integrative Biology and Systems Medicine (IBSE), Robert Bosch Centre for Data Science and AI (RBCDSAI), Wadhwani School of Data Science and Artificial Intelligence (WSAI), IIT Madras, and Aryabhata Data Science and AI Program at THSTI (ADAPT), THSTI.

## AUTHORS’ CONTRIBUTIONS

RT, HS and SB conceived this study; VPG, A, DS and AS pre-processed the data; VPG and AS performed the exploratory data analysis; VPG and RT derived the phenotypic features; VPG, SBT, AS and A performed data and statistical analyses; VPG, A, RT and HS verified the data and compiled the results; VPG, SR, RT and HS interpreted the results; VPG, RT and HS wrote the first draft of the manuscript and VPG, SR, RT and HS critically revised and edited subsequent manuscript drafts; all authors reviewed and approved the manuscript and had full access to the data.

## ETHICS APPROVAL AND CONSENT TO PARTICIPATE

Ethics approvals were obtained from the Institutional Ethics Committees of Translational Health Science and Technology Institute; District Civil Hospital, Gurugram; Safdarjung Hospital, New Delhi (ETHICS/GHG/2014/1.43); Indian Institute of Technology Madras (IEC/2019-03/HS/01/07). Written informed consent was obtained from all study participants enrolled in the GARBH-Ini cohort. All the methods were performed following the relevant guidelines and regulations.

## SUPPLEMENTARY MATERIAL

A supplementary material document is available for this manuscript.

## APPENDIX I Members of the GARBH-Ini Study Group

Translational Health Science and Technology Institute, Faridabad, Haryana, India (Shinjini Bhatnagar, Nitya Wadhwa, Uma Chandra Mouli Natchu, Bhabatosh Das, Pallavi S. Kshetrapal, Shailaja Sopory, Ramachandran Thiruvengadam, Sumit Misra, Dharmendra Sharma, Kanika Sachdeva, Amanpreet Singh, Balakrish G. Nair, Satyajit Rath, Vineeta Bal)

Gurugram Civil Hospital, Haryana, India (Alka Sharma, Sunita Sharma, Umesh Mehta, Brahmdeep Sindhu)

Vardhman Mahavir Medical College & Safdarjung Hospital, New Delhi, India (Pratima Mittal, Rekha Bharti, Harish Chellani, Rani Gera, Jyotsna Suri, Pradeep Debata, Sugandha Arya)

National Institute of Biomedical Genomics, Kalyani, West Bengal, India (Arindam Maitra)

Regional Centre for Biotechnology, Faridabad, Haryana, India (Tushar K. Maiti)

International Centre for Genetic Engineering and Biotechnology, New Delhi, India (Dinakar M. Salunke)

All India Institute of Medical Sciences, New Delhi, India (Nikhil Tandon, Yashdeep Gupta, Alpesh Goyal, Smriti Hari, Aparna Sharma K, Anubhuti Rana)

Maulana Azad Medical College, New Delhi, India (Siddarth Ramji, Anju Garg)

The Ultrasound Lab, Defence Colony, New Delhi, India (Ashok Khurana)

Sitaram Bhartia Institute of Science and Research, New Delhi, India (Reva Tripathi)

President of India’s Secretariat, Government of India (Rakesh Gupta)

Indian Institute of Technology Madras, Chennai, Tamil Nadu, India (Himanshu Sinha, Raghunathan Rengaswamy)

National Science Chair, Science & Engineering Board, Government of India (Partha P. Majumder)

Indian Institute of Science Education and Research, Pune, Maharashtra, India (Vineeta Bal)

Amrita Institute of Medical Sciences & Research Centre, Faridabad, Haryana, India (Pratima Mittal)

Center for Health Research and Development, Society for Applied Studies, New Delhi, India (Uma Chandra Mouli Natchu, Harish Chellani)

Pondicherry Institute of Medical Sciences, Puducherry, India (Ramachandran Thiruvengadam)

