## Supplementary Info for "Phenotypic Characteristics Associated with Preterm Births in the Indian Population"

### **SUPPLEMENTARY INFORMATION**

##### **Hierarchical clustering - optimal number of clusters**

We utilised silhouette scores to determine the optimal number of clusters or assess clustering quality at a particular hierarchical level. This metric is widely employed for evaluating clustering efficacy in diverse unsupervised learning approaches. It offers a data-driven approach to selecting the number of clusters. It considers the distances between data points, clusters, and their centroids to objectively measure how well the data points are grouped within clusters. Its range spans from -1 to 1, with higher values signifying more well-defined clusters. Notably, elevated values tend to correspond with a larger number of clusters, yielding smaller, distinct, and more homogeneous groups. However, given that we aimed to employ statistical testing to identify feature overrepresentation within clusters, larger cluster sizes were desirable. Consequently, we scrutinised silhouette score distributions and cluster sizes across various cluster counts (K clusters) (Figure S4). As a result of this analysis, we determined to prune the hierarchy to achieve 18 clusters, ensuring the creation of adequately large clusters suitable for robust statistical enrichment analysis.

**SUPPLEMENTARY FIGURES**

**Figure S1:** **Distribution of maternal variables across the study cohort.**

Bar plots show the frequency distribution of key maternal phenotypic variables recorded during the study. Each subplot represents a distinct variable, with bars colour-coded to indicate the presence (1 or “yes/present”, orange), absence (0 or “no/absent”, blue), or missing data (NA, green) for that variable. The counts above each bar denote the number of participants in each category. The total number of participants in the dataset was 5232.

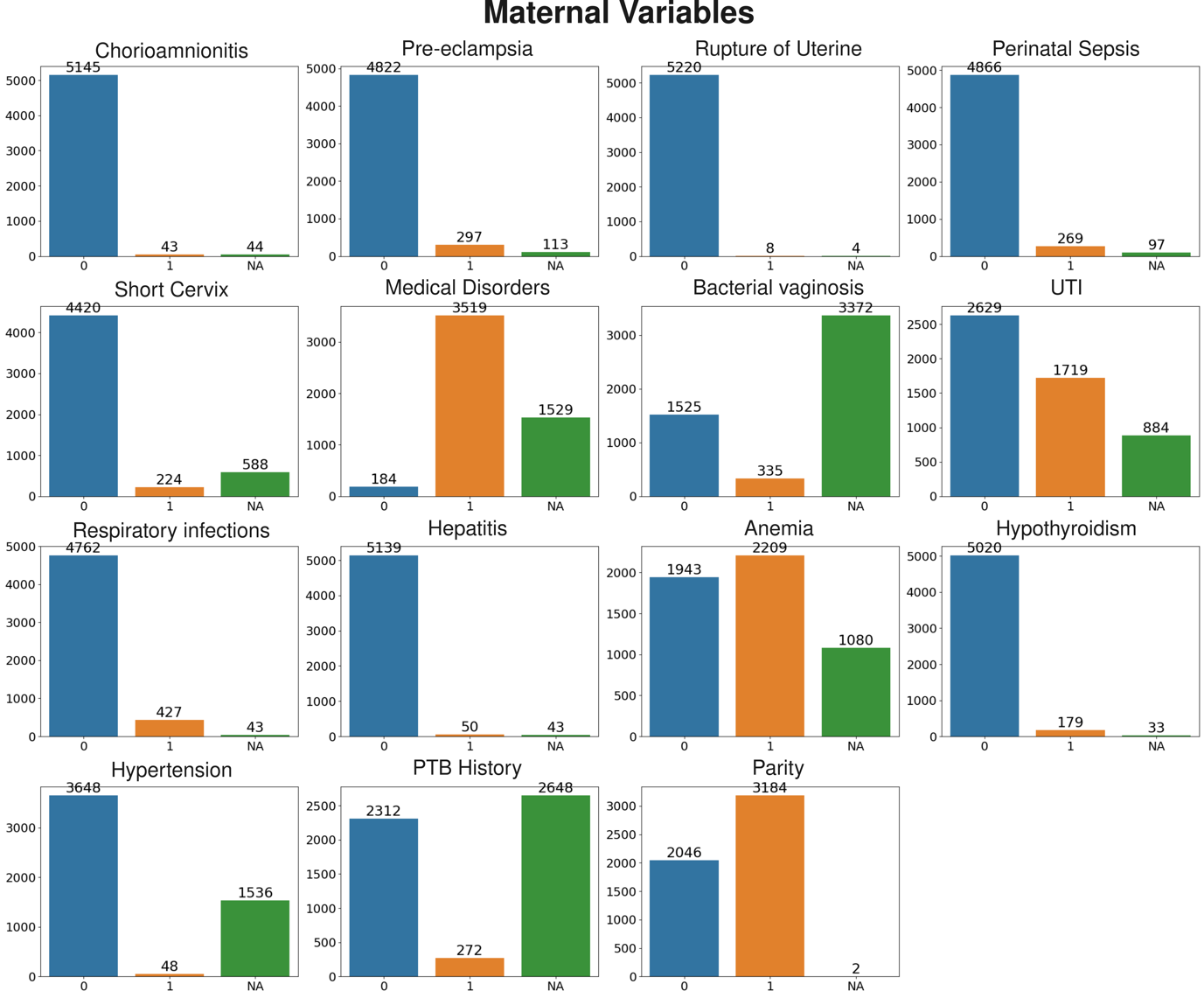

**Figure S2:** **Distribution of foetal, placental, parturition, and other variables across the study cohort.**

Bar plots show the frequency distribution of key maternal phenotypic variables recorded during the study. Each subplot represents a distinct variable, with bars colour-coded to indicate the presence (1 or “yes/present”, orange), absence (0 or “no/absent”, blue), or missing data (NA, green) for that variable. The counts above each bar denote the number of participants in each category. The total number of participants in the dataset was 5232.

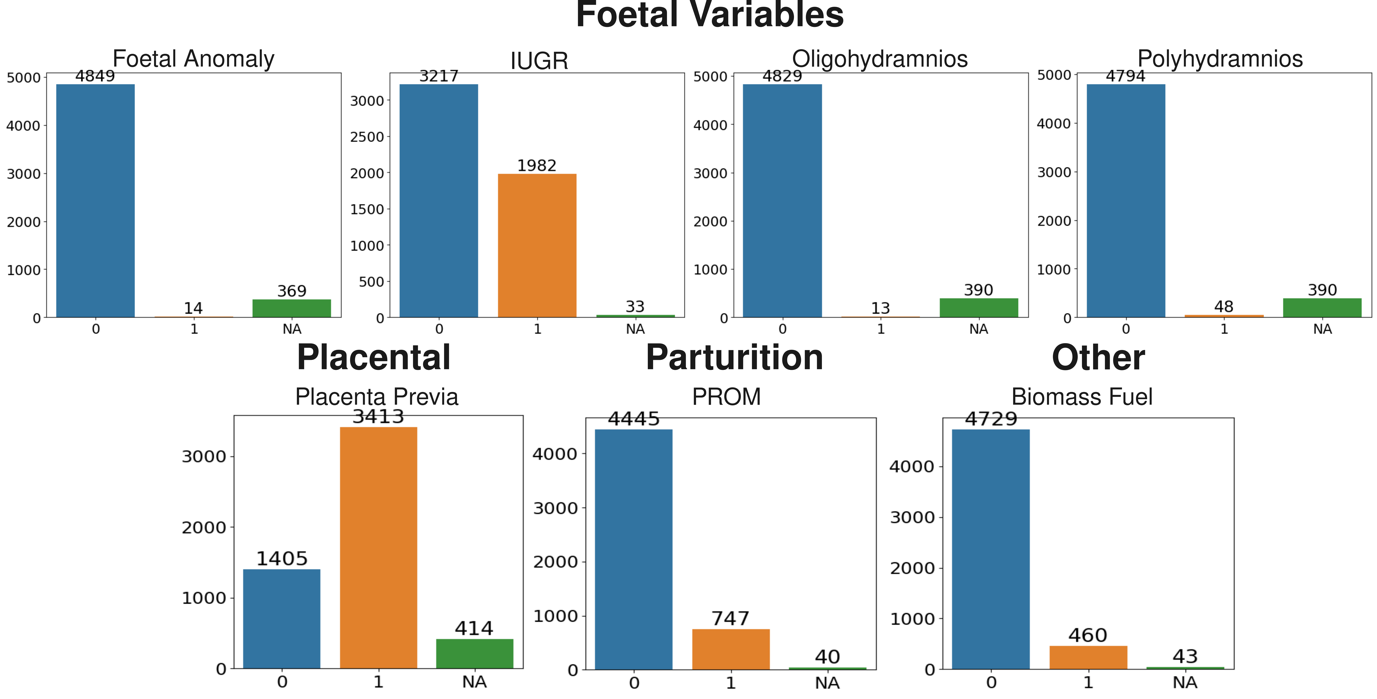

**Figure S3:** **Distribution of Infant variables across the study cohort.**

Bar plots show the frequency distribution of key maternal phenotypic variables recorded during the study. Each subplot represents a distinct variable, with bars colour-coded to indicate the presence (1 or “yes/present”, orange), absence (0 or “no/absent”, blue), or missing data (NA, green) for that variable. The counts above each bar denote the number of participants in each category. The total number of participants in the dataset was 5129.

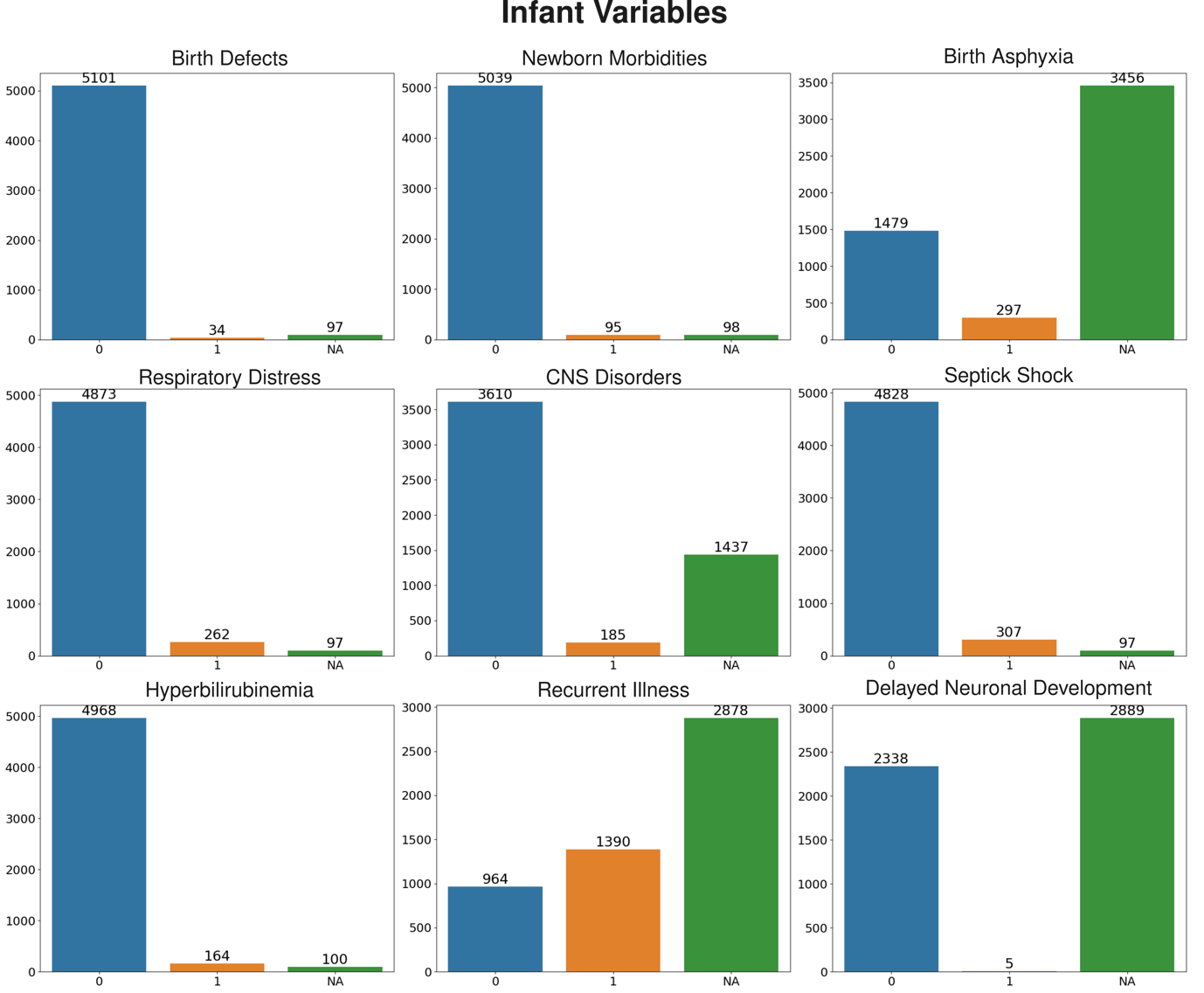

**Figure S4. Distribution of amniotic fluid index (AFI) and cervical length (CL) across PTB and non-PTB participants.**

Histograms show the distribution of two continuous phenotypic variables, AFI (in cm) and CL (in mm), recorded during pregnancy. Participants are colour-coded based on the birth outcome: orange represents preterm birth (1; PTB; N = 609), and blue represents non-PTB (0; N = 4623). The total number of participants included in the analysis was 5232. These distributions illustrate the variability in AFI and CL measurements between PTB and non-PTB groups.

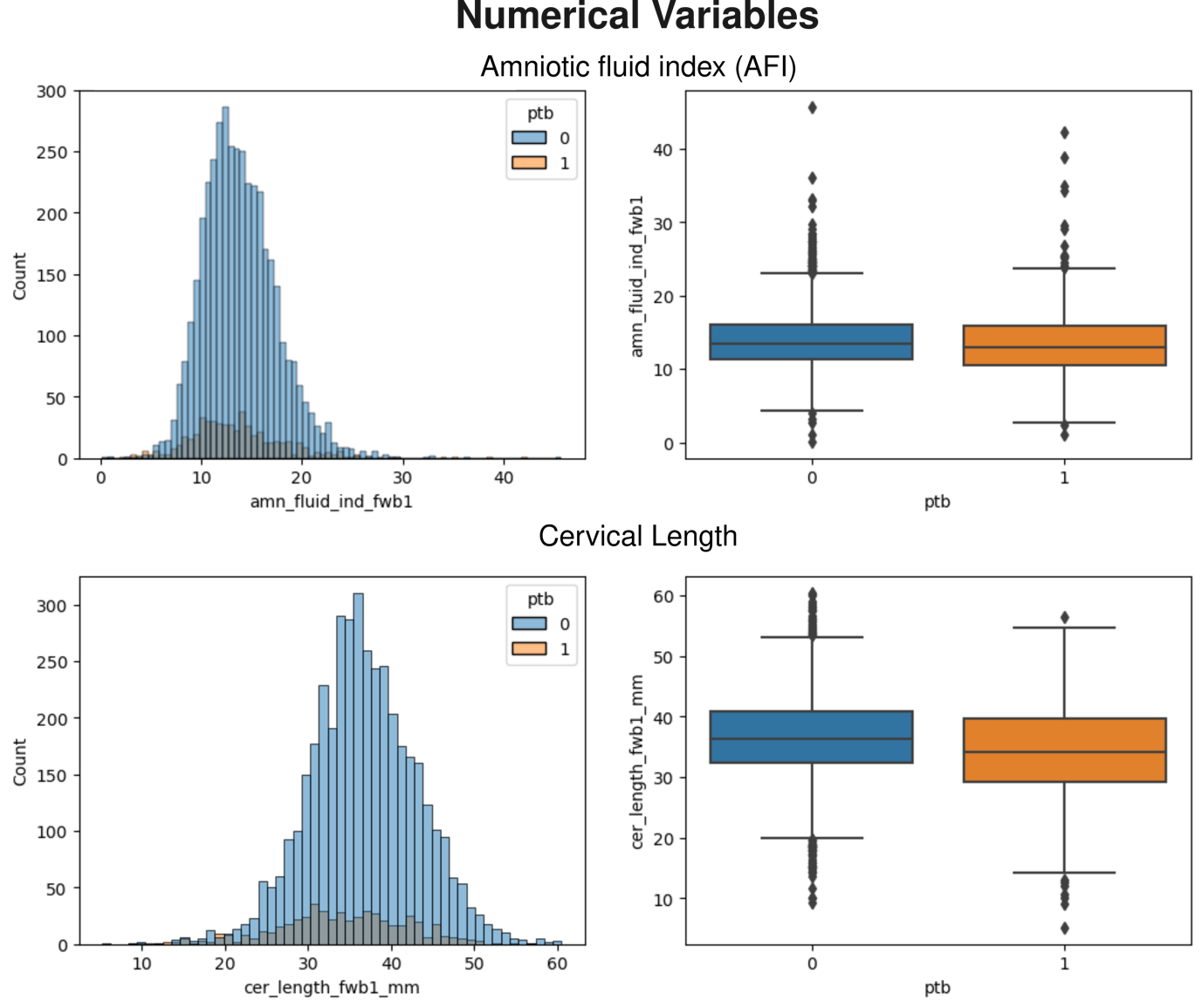

**Figure S5. Evaluation of clustering quality and cluster size distribution for different phenotypic datasets.**

The top row shows the average silhouette scores across 100 random samples for hierarchical clustering on three datasets: Phenotype I (full selected dataset for clustering, N = 3640), Phenotype II – SOL subset (N = 3096), and Phenotype II – CGI subset (N = 111). Higher silhouette scores indicate better-defined clusters. The horizontal red line highlights the silhouette score on the axis corresponding to *K* = 20. The bottom row shows the distribution of cluster sizes for each number of clusters (n) used to cut the dendrogram. The red lines in the violin plots indicate the median cluster size across the 100 samples. The median cluster sizes were 13.82 for the full dataset, 10.56 for the SOL subset, and 1.32 for the CGI subset.

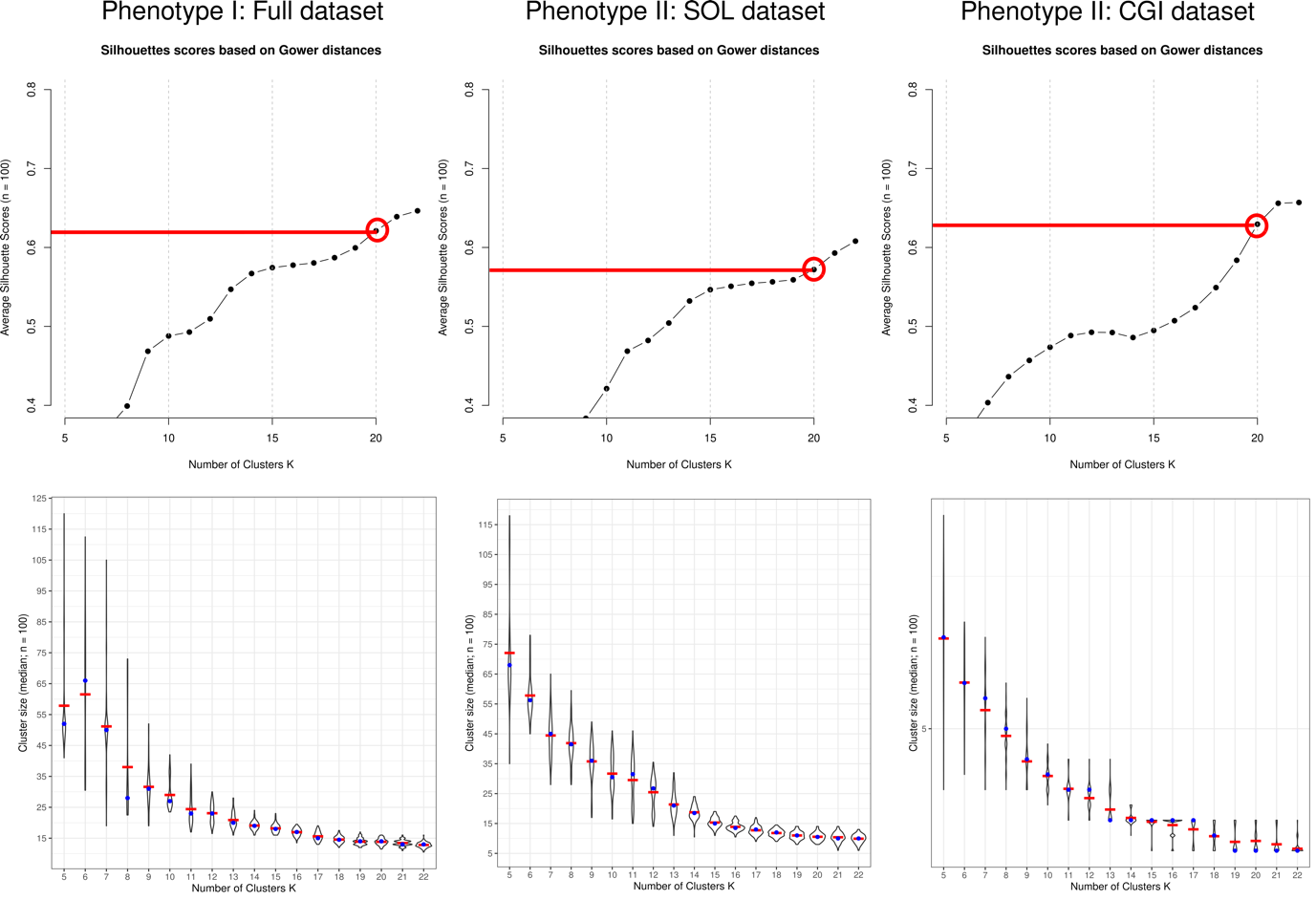

**Figure S6. Distribution of the percentage of ‘Yes’ responses for clinical variables in PTB and non-PTB clusters in Phenotype I.**

This figure displays the distribution of the percentage of cases recorded as ‘Yes’ for various phenotyping variables across clusters derived from the full selected dataset (N = 3640) used in the Phenotype I analysis. The green colour represents the distribution for PTB clusters, while purple represents the non-PTB clusters. These distributions highlight differences in the prevalence of key clinical features between PTB-dominant and non-PTB-dominant clusters. The adjusted P-values for multiple testing are shown on the right side of each distribution.

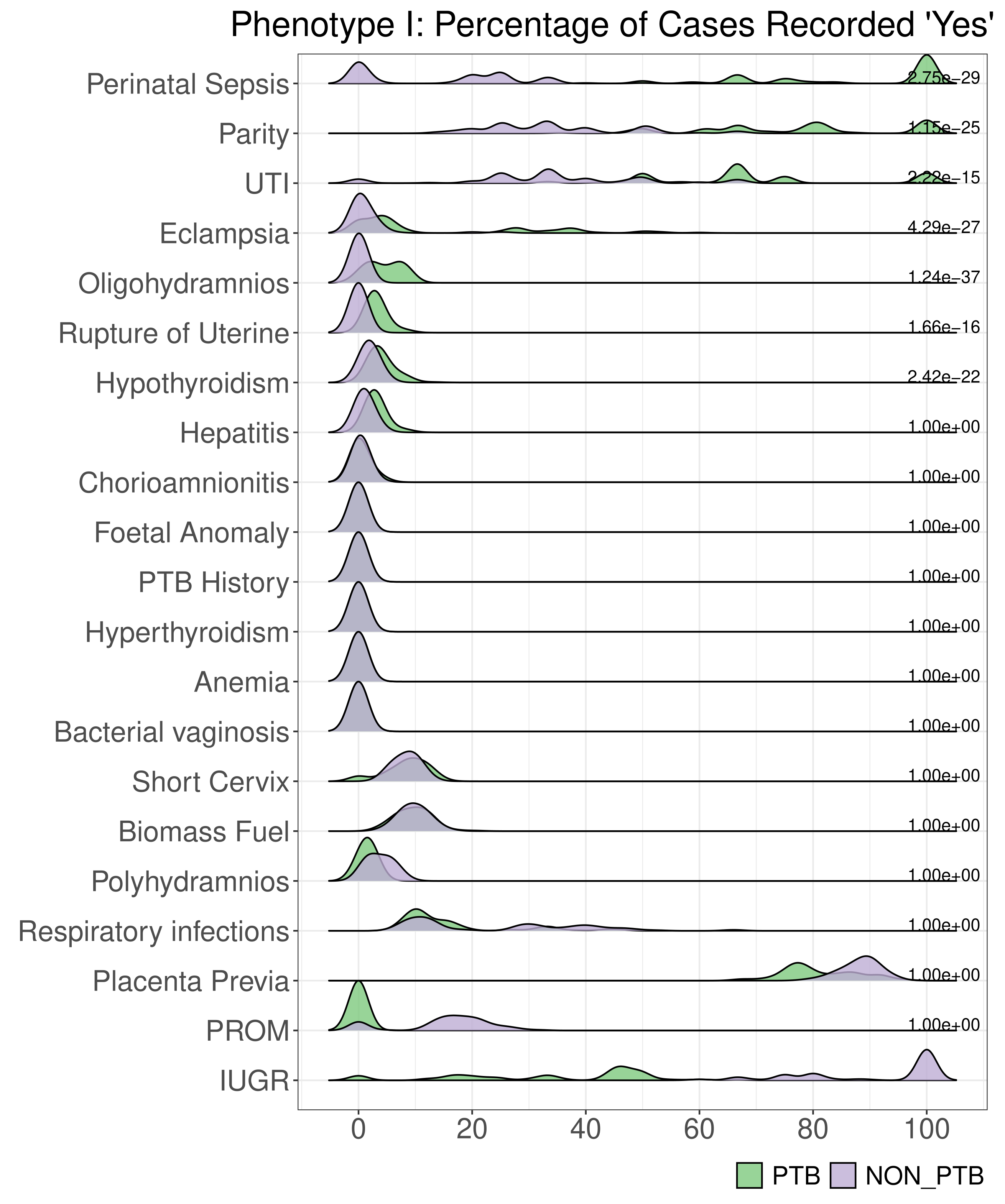

**Figure S7. Distribution of the percentage of ‘Yes’ responses for combinations of phenotypic variables in PTB and non-PTB clusters in Phenotype I.**

This figure shows the distribution of the percentage of cases with a ‘Yes’ response for pairwise combinations of phenotypic variables, stratified by clusters predominantly enriched for preterm birth (PTB, in green) and non-PTB (in purple). These distributions are derived from the full dataset (N = 3640) used in the Phenotype I clustering analysis. The visual comparison highlights differences in the co-occurrence of clinical features between PTB- and non-PTB-dominant clusters. Adjusted p-values from statistical testing (corrected for multiple comparisons) are displayed alongside each variable pair, indicating the significance of observed differences.

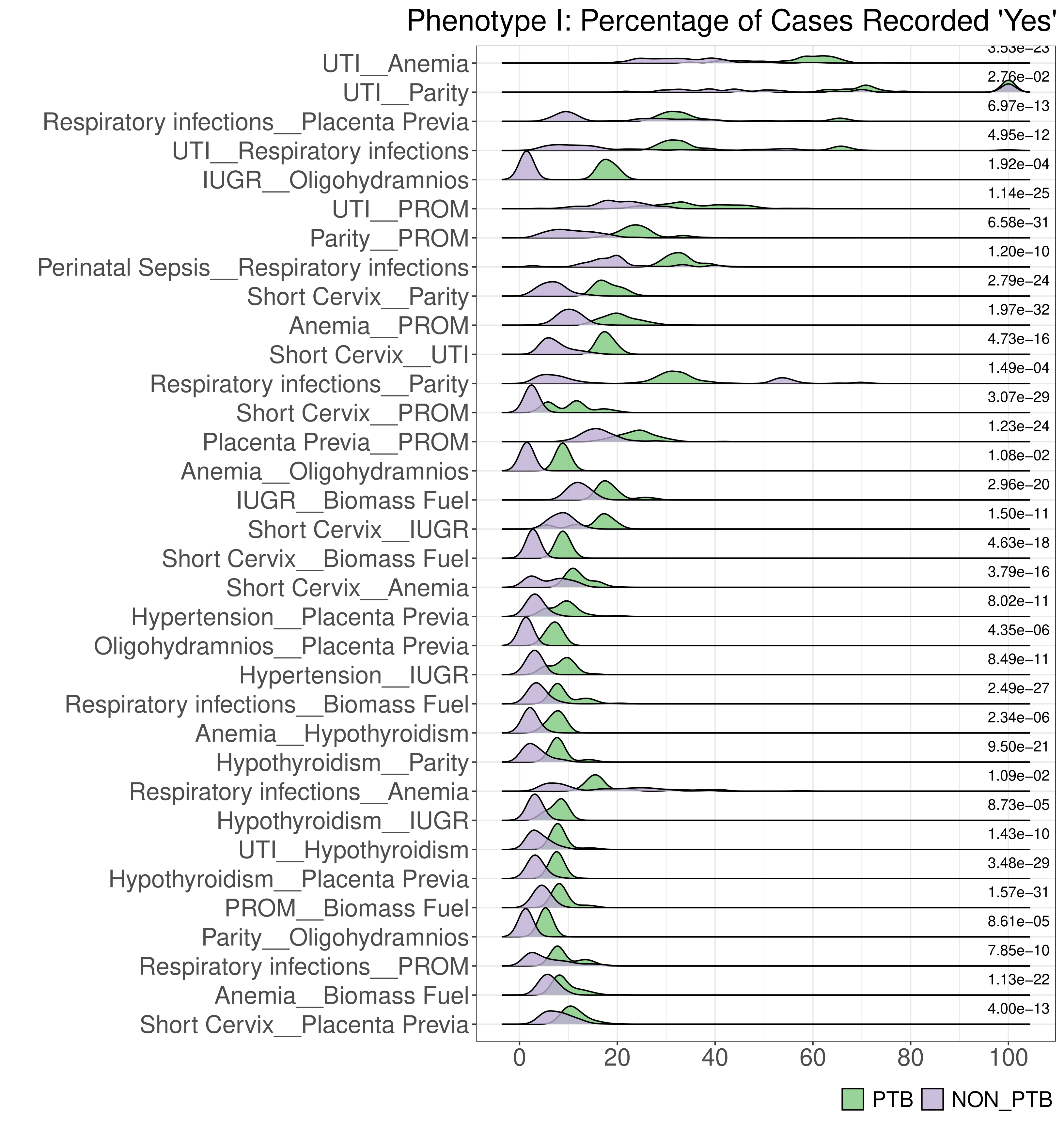

**Table S1:** Baseline characteristics of the participants included and excluded in the study.

| **Sociodemographic characteristics** | **Selected Participants (N = 5231) set median (IQR) or N (%) or mean ± SD** | **Rest of the Participants set (N = 5264) median (IQR) or N (%) or mean ± SD** |
| --- | --- | --- |
| Age (years) | 24 (21, 26) | 24 (21, 26) |
| **BMI at enrolment into the cohort** | | |
| Underweight | 24.90 % | 27.03 % |
| Normal | 58.47 % | 58.54 % |
| Overweight | 12.59 % | 10.31 % |
| Obese | 2.42 % | 1.86 % |
| Haemoglobin (g/dL) | 9.2 (8.6–10.4) | 9.1 (8.6–10.0) |
| Weight (kgs) | 58 | 55 |
| **Socioeconomic status** | | |
| 0 | 0.26 % | 0.47 % |
| 1 | 11.01 % | 12.04 % |
| 2 | 26.34 % | 28.64 % |
| 3 | 38.48 % | 41.62 % |
| 4 | 0.82 % | 0.75 % |
| Undetermined | 23.07 % | 16.43 % |
| **Level of education** | | |
| Illiterate | 16.97 % | 19.87 % |
| Literate or primary school | 8.96 % | 7.92 % |
| Middle school | 16.34 % | 13.29 % |
| High school | 19.02 % | 17.95 % |
| Post-high school diploma | 21.46 % | 21.90 % |
| Graduate | 14.01 % | 14.68 % |
| Post-graduate | 3.21 % | 4.33 % |
| Unknown | 0.00 % | 0.01 % |
| **Occupation** | | |
| Unemployed | 91.35 % | 90.91% |
| Unskilled worker | 3.80 % | 3.91 % |
| Semi-skilled worker | 1.62 % | 1.42 % |
| Skilled worker | 2.46 % | 2.43 % |
| Clerk, shop, farm owner | 0.30 % | 0.28 % |
| Semi-professional | 0.22 % | 0.64 % |
| Professional | 0.21 % | 0.32 % |
| Unknown | 0.00 % | 0.03 % |
| **History of any chronic illnesses** | | |
| Absent | 97.03 % | 97.56 % |
| Present | 2.98 % | 2.43 % |

**Table S2:** Definition of the phenotypic variables used in the study.

|  | **Phenotypic Variables** | **Description** |
| --- | --- | --- |
|  | **Maternal Variables** | |
| 1 | Short Cervix | Cervical length measured by ultrasound shorter than 25 mm |
| 2 | Chorioamnionitis | Diagnosed if the participant had a temperature greater than 38°C and met any of the following conditions: pulse greater than 100 BPM with foul-smelling amniotic fluid, pulse greater than 100 BPM with a foetal heart rate over 160 BPM, or both foul-smelling amniotic fluid and a foetal heart rate over 160 BPM. |
| 3 | Bacterial vaginosis | Diagnosed using Amsel's criteria. BV is diagnosed when at least three out of the following four criteria are met:  Homogenous, thin, greyish-white vaginal discharge that coats the vaginal walls evenly  Vaginal pH greater than 4.5  Presence of clue cells (vaginal epithelial cells with adherent bacteria, obscuring cell borders) on saline wet mount microscopy  Positive "whiff test" (release of a fishy odour when 10% potassium hydroxide is added to a sample of vaginal discharge) |
| 4 | Hepatitis | Diagnosis made by clinician based on physical examination and abnormal laboratory parameters |
| 5 | Respiratory infections | Diagnosis made by clinician based on physical examination |
| 6 | UTI | Diagnosis made by clinician based on presence of fever with burning and/or increased frequency of micturition and/or lower abdominal pain |
| 7 | Pre-eclampsia | Pre-eclampsia was confirmed if the participant had systolic blood pressure ≥ 140 mm/Hg or diastolic blood pressure ≥ 90 mm/Hg, combined with the presence of urine proteins at least 1+ by dipstick test. Eclampsia was confirmed if the participant experienced convulsions and had pre-eclampsia. |
| 8 | Anaemia | Anaemia in pregnancy, as per the Indian Council of Medical Research (ICMR), is defined based on haemoglobin (Hb) concentration: Anaemia: Hb <11 g/dl |
| 9 | Hyperthyroidism | TSH <0.1 mU/L |
| 10 | Hypothyroidism | First trimester: TSH > 2.5 mIU/L |
| 11 | Parity | Number or prior pregnancies that crossed the period of viability (28w of gestation) |
| 12 | Perinatal Sepsis | Perinatal Sepsis was confirmed if any of the following conditions were recorded as positive: antibiotic use, systemic infections, or pneumonia. |
| 13 | PTB History | Previous history of preterm as per history by the participant |
| 14 | Rupture of Uterine | Uterine rupture occurs when the uterine wall tears open. It is more common in individuals attempting a vaginal delivery after a previous caesarean section. Although rare, it is a life-threatening complication that requires immediate medical attention.    Confirmed if there was abdominal pain with bleeding was present, there was a history of caesarean sections, and vaginal bleeding was absent, provided the participant has been pregnant more than once. |
|  | **Foetal Variables** | |
| 15 | IUGR | Estimated foetal weight <10th centile for gestational age as compared against the INTERGROWTH-21st standards |
| 16 | Foetal Anomaly | Structural anomalies impact the developing baby’s body parts, including the heart, lungs, kidneys, limbs, or facial features.  Confirmed if any developmental anomalies or birth defects were recorded either by antenatal ultrasound or birth data |
| 17 | Polyhydramnios | AFI exceeds 24 cm in the third trimester ultrasound examination |
| 18 | Oligohydramnios | AFI less than 5 cm in the third trimester ultrasound examination |
|  | **Placental Variable** | |
| 19 | Placenta Previa | Evidence of the inferior limit spanning across the internal opening of the cervix that connects the uterus to the vagina |
|  | **Parturition Variable** | |
| 20 | PROM | Confirmed if the amniotic sac ruptured prematurely before the onset of labour as evidence by leaking per vaginum during the clinical examination. |
|  | **Other Variable** | |
| 21 | Biomass Fuel | Use of non-LPG, non-electric fuels for cooking |

**Table S3:** Definition of the newborn condition-specific variables.

|  | **Infant Variables** | **Description** |
| --- | --- | --- |
| 1 | Birth Defects | If any defects that can affect the infant’s physical form (such as heart defects, cleft palate, or limb deformities) or functional development (such as metabolic disorders or neural tube defects) |
| 2 | Newborn Morbidities | It is a health complication that can affect infants after birth. It is defined if any of these key variables were recorded as “yes”:   i. Birth Trauma, which indicates whether the infant experienced any physical injury during delivery. ii. Hypoglycaemia refers to low blood sugar levels in the newborn. iii. Meconium Aspiration occurs when a newborn inhales a mixture of meconium and amniotic fluid during delivery. iv. Hypothermia, indicating a dangerously low body temperature in the newborn. |
| 3 | Birth Asphyxia | It is a severe condition in which a newborn receives insufficient oxygen during or immediately after birth, which can lead to serious complications. The presence of certain medical interventions indicates that the newborn experienced this condition. These include IV Fluids, which shows whether intravenous fluids were administered; CPAP, which refers to Continuous Positive Airway Pressure used to assist with breathing; and IMV, which stands for Intermittent Mandatory Ventilation, a method for mechanical ventilation. Additionally, the recording of any central nervous system (CNS) disorders also suggests the occurrence of birth asphyxia. |
| 4 | Respiratory Distress | It refers to a range of conditions affecting newborns that result in difficulty breathing. It is confirmed if any of the following key variables were recorded as “yes”: RDS, which indicates the presence of respiratory distress syndrome; Pneumonia, referring to the diagnosis of pneumonia in the newborn; and Transient Tachypnoea of Newborn, which represents a temporary condition characterised by rapid breathing shortly after birth. |
| 5 | CNS disorders | It refers to conditions affecting the central nervous system in newborns. It is confirmed that either of the two key variables was recorded as “yes”: CNS HIE, which indicates whether the infant has experienced hypoxic-ischemic encephalopathy, and CNS SEI, which denotes the occurrence of seizures. |
| 6 | Septic Shock | it is a severe condition that occurs when an infection leads to dangerously low blood pressure and insufficient blood flow to the organs, which can be life-threatening. It is confirmed if either of these two key variables were recorded as “yes”: the presence of systemic infection and the administration of intravenous fluids. |
| 7 | Liver Disorders | It is a condition that can lead to jaundice in newborns. It is confirmed if hyperbilirubinemia is recorded in newborns. |
| 8 | Recurrent Illness | It is a condition that is confirmed if the baby was taken to a doctor for reasons other than immunisation if the baby fell ill and required medical consultation more than once, or if the baby has been hospitalised at any point since birth. |
| 9 | Delayed neuronal development | A condition characterised by an infant’s delay in reaching important developmental milestones related to brain and sensory processing. It is confirmed if any of the following is recorded as “yes”: infant’s ability to smile at its mother, turn its head toward sounds, make sounds, and lift its head while lying on its stomach. |

**Table S4.** **Phenotypic variables selected for clustering analysis based on data completeness and prevalence.** This table lists the subset of phenotypic variables used for clustering analysis after applying inclusion criteria: variables with <20% missing data and ≥5% of cases recorded as “Yes” in the complete dataset (N = 5232). Variables are grouped by category (Maternal, Foetal, Placental, Other), and the distribution of responses is shown as the percentage of “Yes,” “No,” and “NA” (missing) values for each variable.

| **Variable Class** | **Phenotypic Variables** | **No %** | **Yes %** | **NA %** |
| --- | --- | --- | --- | --- |
| Maternal | Pre-eclampsia | 92.16 | 5.68 | 2.16 |
| Maternal | Perinatal Sepsis | 93.00 | 5.14 | 1.85 |
| Maternal | UTI | 50.25 | 32.86 | 16.90 |
| Maternal | Respiratory infections | 91.02 | 8.16 | 0.82 |
| Maternal | Parity | 39.11 | 60.86 | 0.04 |
| Foetal | IUGR | 61.49 | 37.88 | 0.63 |
| Placental | Placenta Previa | 26.85 | 65.23 | 7.91 |
| Other | Biomass Fuel | 90.39 | 8.79 | 0.82 |

**Table S5:** **Neonatal characteristics associated with phenotypic variables in PTB and non-PTB cases.** This table summarises the gestational age (GA), birth weight (BW), birth length (BL), and head circumference (HC) at birth, along with the frequency of Neonatal Intensive Care Unit (NICU) admissions and Early Neonatal Deaths (END), for each phenotypic condition observed in the study cohort (N = 5232). For each condition, values are reported separately for preterm birth (PTB) and non-preterm birth (non-PTB) groups. Continuous variables (GA, BW, BL, HC) are presented as mean (standard deviation), while NICU and END outcomes are shown as counts (percentages).

| **Phenotypic variables** | **Class** | **Mean (Standard deviation)** | | | | | **Counts (Percentage)** | |
| --- | --- | --- | --- | --- | --- | --- | --- | --- |
|  |  | **GA (Weeks)** | **BW (kg)** | **BL (cm)** | **HC (cm)** | **NICU** | | **END** |
| Chorioamnionitis | PTB | 33.5 (1.76) | 1.89 (0.39) | 44.2 (3.83) | 30.85 (1.9) | 3 (50) | | 1 (17) |
|  | Non-PTB | 38.68 (1.2) | 2.71 (0.43) | 48.3 (1.6) | 33.27 (1.04) | 1 (2.7) | | 0 (0) |
| Eclampsia | PTB | 34.26 (1.78) | 1.97 (0.59) | 44.46 (3.41) | 31.04 (1.73) | 36 (62.07) | | 5 (9) |
|  | Non-PTB | 38.95 (1.27) | 2.72 (0.50) | 48.19 (2.42) | 33.02 (1.52) | 53 (22.18) | | 2 (1) |
| Rupture of Uterine | PTB | 32 (4.24) | 1.69 (0.69) | 46.3 (3.25) | 28.75 (2.47) | 2 (100) | | 1 (50) |
|  | Non-PTB | 38.5 (1.38) | 2.68 (0.35) | 47.27 (2.11) | 32.88 (1.01) | 0 (0) | | 0 (0) |
| Perinatal Sepsis | PTB | 33.18 (1.93) | 1.74 (0.44) | 43.34 (3.53) | 30.4 (2.07) | 101 (100) | | 13 (13) |
|  | Non-PTB | 39.23 (1.27) | 2.64 (0.45) | 47.96 (2.38) | 33.21 (1.49) | 165 (98.21) | | 19 (11) |
| Short Cervix | PTB | 33.95 (1.85) | 2.04 (0.42) | 44.27 (3.23) | 30.97 (1.51) | 27 (40.91) | | 2 (3) |
|  | Non-PTB | 38.59 (1.17) | 2.75 (0.44) | 47.5 (2.53) | 32.8 (1.16) | 14 (8.86) | | 1 (1) |
| Bacterial vaginosis | PTB | 34.8 (1.51) | 2.24 (0.44) | 45.12 (4.21) | 32.01 (1.16) | 11 (31.43) | | 2 (6) |
|  | Non-PTB | 39.09 (1.2) | 2.86 (0.42) | 48.21 (2.23) | 33.34 (1.3) | 17 (5.67) | | 4 (1) |
| UTI | PTB | 34.93 (1.48) | 2.20 (0.43) | 45.7 (2.76) | 31.72 (1.49) | 58 (29.44) | | 4 (2) |
|  | Non-PTB | 38.91 (1.15) | 2.82 (0.41) | 48.19 (2.05) | 33.24 (1.27) | 96 (6.31) | | 11 (1) |
| Respiratory infections | PTB | 34.76 (1.6) | 2.16 (0.50) | 45.17 (3.08) | 31.36 (1.7) | 16 (32) | | 2 (4) |
|  | Non-PTB | 38.8 (1.14) | 2.82 (0.41) | 48.29 (1.87) | 33.2 (1.2) | 36 (9.55) | | 2 (1) |
| Hepatitis | PTB | 35.22 (1.09) | 2.32 (0.19) | 46.24 (2.41) | 31.48 (1.4) | 2 (22.22) | | 0 (0) |
|  | Non-PTB | 38.85 (1.26) | 2.64 (0.34) | 47.95 (1.87) | 32.85 (1.13) | 4 (9.76) | | 1 (2) |
| Anaemia | PTB | 34.87 (1.52) | 2.21 (0.42) | 45.85 (2.64) | 31.78 (1.56) | 71 (25.63) | | 5 (2) |
|  | Non-PTB | 38.96 (1.17) | 2.84 (0.42) | 48.3 (2.04) | 33.27 (1.27) | 116 (6) | | 16 (1) |
| Hypothyroidism | PTB | 35.26 (1.06) | 2.33 (0.51) | 46.26 (3.3) | 32.14 (1.45) | 13 (41.94) | | 0 (0) |
|  | Non-PTB | 38.91 (1.12) | 2.90 (0.46) | 48.4 (2.08) | 33.37 (1.32) | 16 (10.81) | | 1 (1) |
| Hypertension | PTB | 34.08 (1.85) | 2.07 (0.88) | 45.19 (3.97) | 32.4 (1.65) | 10 (76.92) | | 0 (0) |
|  | Non-PTB | 38.6 (1.12) | 2.62 (0.47) | 48.04 (2.57) | 33.01 (1.17) | 8 (22.86) | | 2 (6) |
| PTB History | PTB | 34.37 (1.87) | 2.20 (0.48) | 45.89 (3.22) | 31.8 (1.86) | 29 (39.73) | | 2 (3) |
|  | Non-PTB | 38.54 (1.09) | 2.78 (0.40) | 48.12 (2.05) | 33.18 (1.32) | 12 (6.03) | | 0 (0) |
| Parity | PTB | 34.82 (1.58) | 2.23 (0.48) | 45.82 (2.95) | 31.75 (1.68) | 114 (30) | | 9 (2) |
|  | Non-PTB | 38.87 (1.13) | 2.87 (0.41) | 48.3 (2.1) | 33.28 (1.22) | 177 (6.31) | | 18 (1) |
| Foetal Anomaly | PTB | 32.75 (0.96) | 1.70 (0.44) | 43 (NA) | 31 (NA) | 4 (100) | | 3 (75) |
|  | Non-PTB | 38.9 (1.37) | 2.91 (0.84) | 48.23 (1.39) | 33.14 (0.62) | 3 (30) | | 3 (30) |
| IUGR | PTB | 34.99 (1.41) | 1.85 (0.36) | 44.42 (2.89) | 31.13 (1.57) | 68 (41.72) | | 6 (4) |
|  | Non-PTB | 39.02 (1.16) | 2.48 (0.26) | 47.43 (2.07) | 32.73 (1.19) | 179 (9.84) | | 13 (1) |
| Oligohydramnios | PTB | 32.9 (2.18) | 1.76 (0.54) | 42.62 (3.26) | 29.68 (1.91) | 7 (70) | | 1 (10) |
|  | Non-PTB | 38.33 (1.15) | 2.59 (0.36) | 46.7 (2.34) | 31.93 (2.53) | 0 (0) | | 0 (0) |
| Polyhydramnios | PTB | 33.77 (1.88) | 1.96 (0.39) | 45.58 (2.45) | 31.61 (1.31) | 5 (38.46) | | 1 (8) |
|  | Non-PTB | 39.09 (1.25) | 3.04 (0.47) | 48.92 (2.39) | 34.09 (1.55) | 7 (20) | | 1 (3) |
| Placenta Previa | PTB | 34.91 (1.37) | 2.20 (0.41) | 45.88 (2.85) | 31.83 (1.54) | 101 (28.37) | | 12 (3) |
|  | Non-PTB | 38.97 (1.17) | 2.83 (0.41) | 48.31 (2.1) | 33.3 (1.25) | 208 (6.8) | | 23 (1) |
| PROM | PTB | 34.39 (1.89) | 2.14 (0.50) | 45.63 (3.15) | 31.36 (1.69) | 47 (35.61) | | 2 (2) |
|  | Non-PTB | 38.76 (1.19) | 2.81 (0.40) | 48.32 (2.04) | 33.19 (1.21) | 52 (8.46) | | 5 (1) |

**Table S6. Pairwise associations of phenotypic variables in the complete dataset (N = 5232; PTB = 609, Non-PTB = 4656)**. This table presents the co-occurrence of phenotypic conditions stratified by PTB and non-PTB cases. For each primary variable (leftmost column), the percentage of participants with co-occurring conditions is shown in the third column. Values highlighted in bold indicate conditions that are at least twice as prevalent in PTB cases compared to non-PTB cases.

| **Phenotypic Variables** | **Class (N)** | **Associated Conditions (%)** |
| --- | --- | --- |
| Anaemia | PTB (277) | Placenta Previa (67.51), Parity (67.15), UTI (35.74), IUGR (25.27), PROM (23.47), **Perinatal Sepsis (14.8)**, **PTB History (11.55),** **Short Cervix (10.11)**, Respiratory infections (10.11), Pre-eclampsia (5.78), Bacterial vaginosis (5.78), Hypothyroidism (3.61), Hepatitis (2.17), Oligohydramnios (1.44), Foetal Anomaly (1.08), Polyhydramnios (1.08), Hypertension (1.08), Chorioamnionitis (0.36), Rupture of Uterine (0.36) |
|  | Non-PTB (1932) | Placenta Previa (79.45), Parity (62.06), IUGR (39.34), UTI (34.06), PROM (15.17), Respiratory infections (9.06), Bacterial vaginosis (6.47), Pre-eclampsia (4.61), **PTB History (4.4)**, **Perinatal Sepsis (3.47)**, **Short Cervix (3.31)**, Hypothyroidism (2.48), Hepatitis (1.35), Polyhydramnios (0.93), Hypertension (0.57), Chorioamnionitis (0.47), Foetal Anomaly (0.31), Rupture of Uterine (0.1), Oligohydramnios (0.05) |
| Bacterial vaginosis | PTB (35) | Placenta Previa (77.14), Parity (57.14), Anaemia (45.71), UTI (37.14), IUGR (20), **PTB History (17.14)**, PROM (17.14), **Perinatal Sepsis (14.29)**, **Pre-eclampsia** (8.57), Respiratory infections (8.57), Short Cervix (5.71), **Hypothyroidism (5.71)**, **Hypertension (5.71)**, Foetal Anomaly (2.86), **Oligohydramnios (2.86)**, **Polyhydramnios (2.86)** |
|  | Non-PTB (300) | Placenta Previa (87.67), Parity (54.33), Anaemia (41.67), UTI (40.67), IUGR (38), PROM (13.33), Respiratory infections (6), **PTB History (4.33)**, **Perinatal Sepsis (4)**, Short Cervix (3.33), **Pre-eclampsia (3)**, **Hypothyroidism (1.67)**, Chorioamnionitis (1.33), Hepatitis (1.33), **Hypertension (1.33)**, **Oligohydramnios (0.67)**, Rupture of Uterine (0.33), **Polyhydramnios (0.33)** |
| Chorioamnionitis | PTB (6) | Placenta Previa (50), Pre-eclampsia (33.33), IUGR (33.33), Parity (33.33), Perinatal Sepsis (16.67), Anaemia (16.67), UTI (16.67) |
|  | Non-PTB (37) | Placenta Previa (70.27), Parity (67.57), IUGR (45.95), UTI (29.73), Anaemia (24.32), PROM (10.81), Bacterial vaginosis (10.81), Respiratory infections (10.81), PTB History (8.11), Pre-eclampsia (5.41), Hypothyroidism (2.7) |
| Pre-eclampsia | PTB (58) | Parity (58.62), Placenta Previa (56.9), IUGR (44.83), Anaemia (27.59), UTI (18.97), **PTB History (15.52)**, **Perinatal Sepsis (13.79)**, PROM (13.79), Hypertension (13.79), Short Cervix (6.9), Hypothyroidism (6.9), Bacterial vaginosis (5.17), Respiratory infections (5.17), Chorioamnionitis (3.45), Foetal Anomaly (1.72), Oligohydramnios (1.72) |
|  | Non-PTB (239) | Placenta Previa (79.5), Parity (60.25), IUGR (51.05), Anaemia (37.24), UTI (33.05), PROM (12.97), Respiratory infections (11.72), Hypertension (10.46), **PTB History (6.28)**, Hypothyroidism (5.02), **Perinatal Sepsis (3.77)**, Short Cervix (3.77), Bacterial vaginosis (3.77), Polyhydramnios (2.09), Rupture of Uterine (1.26), Chorioamnionitis (0.84), Hepatitis (0.84), Foetal Anomaly (0.42) |
| Foetal Anomaly | PTB (4) | Perinatal Sepsis (100), Placenta Previa (75), Anaemia (75), Parity (75), Short Cervix (50), Polyhydramnios (50), IUGR (50), Pre-eclampsia (25), PROM (25), Bacterial vaginosis (25), UTI (25), Respiratory infections (25) |
|  | Non-PTB (10) | Perinatal Sepsis (100), Placenta Previa (75), Anaemia (75), Parity (75), Short Cervix (50), Polyhydramnios (50), IUGR (50), Pre-eclampsia (25), PROM (25), Bacterial vaginosis (25), UTI (25), Respiratory infections (25) |
| Hepatitis | PTB (9) | Placenta Previa (80), Anaemia (60), UTI (50), Parity (50), IUGR (30), Perinatal Sepsis (20), **Pre-eclampsia (10)**, Polyhydramnios (10), Hypothyroidism (10), Respiratory infections (10), Hepatitis (10), Hypertension (10) |
|  | Non-PTB (41) | Placenta Previa (68.29), Parity (65.85), Anaemia (63.41), UTI (63.41), IUGR (51.22), Respiratory infections (26.83), PROM (9.76), Bacterial vaginosis (9.76), PTB History (7.32), **Pre-eclampsia (4.88)**, Perinatal Sepsis (2.44), Foetal Anomaly (2.44), Hypothyroidism (2.44), Hypertension (2.44) |
| Hypertension | PTB (13) | Placenta Previa (61.54), Pre-eclampsia (61.54), Parity (61.54), IUGR (38.46), **PROM (30.77)**, Anaemia (23.08), PTB History (15.38), Bacterial vaginosis (15.38), Hypothyroidism (15.38), Perinatal Sepsis (7.69), Short Cervix (7.69), UTI (7.69), Respiratory infections (7.69) |
|  | Non-PTB (35) | Placenta Previa (85.71), Parity (74.29), Pre-eclampsia (71.43), IUGR (54.29), Anaemia (31.43), UTI (31.43), PTB History (14.29), Respiratory infections (14.29), **PROM (11.43)**, Bacterial vaginosis (11.43), Hypothyroidism (11.43), Perinatal Sepsis (8.57), Foetal Anomaly (2.86), Rupture of Uterine (2.86), Polyhydramnios (2.86), Hepatitis (2.86) |
| Hypothyroidism | PTB (31) | Parity (70.97), Placenta Previa (32.26), Anaemia (32.26), UTI (32.26), IUGR (25.81), PROM (22.58), **PTB History (19.35)**, **Perinatal Sepsis (12.9)**, Pre-eclampsia (12.9), **Short Cervix (12.9)**, **Respiratory infections (12.9)**, Bacterial vaginosis (6.45), **Hypertension (6.45)**, Oligohydramnios (3.23) |
|  | Non-PTB (148) | Parity (70.27), Placenta Previa (46.62), IUGR (34.46), Anaemia (32.43), UTI (27.7), PROM (14.19), Pre-eclampsia (8.11), **PTB History (6.76)**, **Respiratory infections (5.41)**, **Perinatal Sepsis (4.05)**, Bacterial vaginosis (3.38), Polyhydramnios (2.7), Hypertension (2.7), **Short Cervix (2.03)**, Chorioamnionitis (0.68), Foetal Anomaly (0.68), Hepatitis (0.68) |
| IUGR | PTB (163) | Placenta Previa (65.03), Parity (52.76), Anaemia (42.94), UTI (36.81), **Perinatal Sepsis (24.54)**, PROM (19.63), pre-eclampsia (15.95), **Short Cervix (10.43)**, **PTB History (9.2)**, Respiratory infections (9.2), Hypothyroidism (4.91), Bacterial vaginosis (4.29), **Hypertension (3.07)**, Oligohydramnios (2.45), **Polyhydramnios (2.45)**, Chorioamnionitis (1.23), **Foetal Anomaly (1.23),** Hepatitis (1.23) |
|  | Non-PTB (1819) | Placenta Previa (67.62), Parity (53.38), Anaemia (41.78), UTI (33.15), PROM (12.81), Respiratory infections (7.59), Pre-eclampsia (6.71), Bacterial vaginosis (6.27), **Perinatal Sepsis (5.44)**, **PTB History (3.85)**, **Short Cervix (3.74)**, Hypothyroidism (2.8), Hepatitis (1.15), **Hypertension (1.04)**, Chorioamnionitis (0.93), **Polyhydramnios (0.38)**, Rupture of Uterine (0.22), **Foetal Anomaly (0.16)**, Oligohydramnios (0.11) |
| Oligohydramnios | PTB (10) | Placenta Previa (60), Parity (50), Perinatal Sepsis (40), Short Cervix (40), IUGR (40), Anaemia (40), PROM (30), UTI (30), PTB History (10), Pre-eclampsia (10), Bacterial vaginosis (10), Hypothyroidism (10) |
|  | Non-PTB (3) | Placenta Previa (100), Parity (100), IUGR (66.67), Bacterial vaginosis (66.67), PROM (33.33), Anaemia (33.33) |
| PROM | PTB (132) | Placenta Previa (58.33), Parity (52.27), Anaemia (49.24), UTI (34.85), **Perinatal Sepsis (24.24)**, IUGR (24.24), **Short Cervix (15.91)**, Respiratory infections (9.85), PTB History (7.58), Pre-eclampsia (6.06), Hypothyroidism (5.3), Bacterial vaginosis (4.55), Polyhydramnios (3.03), Hypertension (3.03), **Oligohydramnios (2.27)**, Hepatitis (1.52), Foetal Anomaly (0.76), Rupture of Uterine (0.76) |
|  | Non-PTB (615) | Placenta Previa (69.43), Parity (53.82), Anaemia (47.64), IUGR (37.89), UTI (34.15), Respiratory infections (8.62), Bacterial vaginosis (6.5), Pre-eclampsia (5.04), PTB History (4.55), **Perinatal Sepsis (4.07)**, Hypothyroidism (3.41), **Short Cervix (2.76)**, Polyhydramnios (1.14), Chorioamnionitis (0.65), Hepatitis (0.65), Hypertension (0.65), Rupture of Uterine (0.16), **Oligohydramnios (0.16)**, Foetal Anomaly (NA) |
| PTB History | PTB (73) | Parity (100), Placenta Previa (58.9), Anaemia (43.84), UTI (34.25), **Perinatal Sepsis (21.92)**, IUGR (20.55), PROM (13.7), Pre-eclampsia (12.33), **Short Cervix (8.22)**, Bacterial vaginosis (8.22), Hypothyroidism (8.22), Respiratory infections (5.48), Polyhydramnios (2.74), Hepatitis (2.74), Hypertension (2.74), **Oligohydramnios (1.37)** |
|  | Non-PTB (199) | Parity (100), Placenta Previa (67.84), Anaemia (42.71), IUGR (35.18), UTI (31.16), PROM (14.07), Respiratory infections (14.07), Pre-eclampsia (7.54), Bacterial vaginosis (6.53), Hypothyroidism (5.03), **Short Cervix (4.02)**, Hypertension (2.51), **Perinatal Sepsis (2.01)**, Chorioamnionitis (1.51), Polyhydramnios (1.51), Hepatitis (1.51), Rupture of Uterine (0.5) |
| Parity | PTB (380) | Placenta Previa (61.32), Anaemia (48.95), UTI (32.63), IUGR (22.63), **PTB History (19.21)**, PROM (18.16), **Perinatal Sepsis (15.26)**, Pre-eclampsia (8.95), **Short Cervix (8.95)**, Respiratory infections (8.42), Hypothyroidism (5.79), Bacterial vaginosis (5.26), Hepatitis (2.37), Hypertension (2.11), Polyhydramnios (1.58), **Oligohydramnios (1.32)**, Foetal Anomaly (0.79), Chorioamnionitis (0.53), Rupture of Uterine (0.53) |
|  | Non-PTB (2804) | Placenta Previa (67.69), Anaemia (42.76), IUGR (34.63), UTI (32.99), PROM (11.8), Respiratory infections (7.74), **PTB History (7.1)**, Bacterial vaginosis (5.81), Pre-eclampsia (5.14), Hypothyroidism (3.71), **Short Cervix (3.17)**, **Perinatal Sepsis (2.82)**, Hepatitis (0.96), Hypertension (0.93), Chorioamnionitis (0.89), Polyhydramnios (0.75), Rupture of Uterine (0.21), Foetal Anomaly (0.18), **Oligohydramnios (0.11**) |
| Perinatal Sepsis | PTB (101) | Parity (57.43), Placenta Previa (52.48), Anaemia (40.59), IUGR (39.6), UTI (36.63), **PROM (31.68)**, **Short Cervix (16.83)**, **PTB History (15.84)**, Respiratory infections (10.89), Pre-eclampsia (7.92), Bacterial vaginosis (4.95), **Foetal Anomaly (3.96)**, **Oligohydramnios (3.96)**, Hypothyroidism (3.96), Polyhydramnios (2.97), **Rupture of Uterine (1.98)**, Hepatitis (1.98), Chorioamnionitis (0.99), Hypertension (0.99) |
|  | Non-PTB (168) | Placenta Previa (64.29), IUGR (58.93), Parity (47.02), Anaemia (39.88), UTI (30.95), **PROM (14.88)**, Respiratory infections (11.9), Bacterial vaginosis (7.14), Pre-eclampsia (5.36), **Short Cervix (4.17)**, Hypothyroidism (3.57), **PTB History (2.38)**, Hypertension (1.79), **Foetal Anomaly (1.19)**, Polyhydramnios (1.19), Hepatitis (0.6) |
| Placenta Previa | PTB (356) | Parity (65.45), Anaemia (52.53), UTI (39.04), IUGR (29.78), PROM (21.63), **Perinatal Sepsis (14.89)**, **PTB History (12.08)**, **Short Cervix (10.96)**, Respiratory infections (9.55), Pre-eclampsia (9.27), Bacterial vaginosis (7.58), Hypothyroidism (2.81), Polyhydramnios (2.25), Hypertension (2.25), **Oligohydramnios (1.69)**, Hepatitis (1.4), Chorioamnionitis (0.84), Foetal Anomaly (0.84), Rupture of Uterine (0.28) |
|  | Non-PTB (3057) | Parity (62.09), Anaemia (50.21), IUGR (40.24), UTI (35.85), PROM (13.97), Respiratory infections (8.8), Bacterial vaginosis (8.6), Pre-eclampsia (6.22), **PTB History (4.42)**, **Perinatal Sepsis (3.53)**, **Short Cervix (2.88)**, Hypothyroidism (2.26), Polyhydramnios (0.98), Hypertension (0.98), Hepatitis (0.92), Chorioamnionitis (0.85), Foetal Anomaly (0.26), Rupture of Uterine (0.13), **Oligohydramnios (0.1)** |
| Polyhydramnios | PTB (13) | Placenta Previa (61.54), **Short Cervix (53.85)**, Parity (46.15), PROM (30.77), IUGR (30.77), **Perinatal Sepsis (23.08)**, Anaemia (23.08), PTB History (15.38), **Foetal Anomaly (15.38)**, Respiratory infections (15.38), **Bacterial vaginosis (7.69)**, UTI (7.69) |
|  | Non-PTB (35) | Placenta Previa (85.71), Parity (60), Anaemia (51.43), UTI (31.43), PROM (20), IUGR (20), Eclampsia (14.29), Respiratory infections (14.29), **Short Cervix (11.43)**, Hypothyroidism (11.43), PTB History (8.57), **Perinatal Sepsis (5.71)**, **Foetal Anomaly (2.86)**, **Bacterial vaginosis (2.86)**, Hypertension (2.86) |
| Respiratory infections | PTB (50) | Placenta Previa (68), Parity (64), Anaemia (56), UTI (38), IUGR (30), PROM (26), **Perinatal Sepsis (22)**, **Short Cervix (10)**, PTB History (8), **Hypothyroidism (8)**, Eclampsia (6), Bacterial vaginosis (6), **Hepatitis (6)**, Polyhydramnios (4), **Foetal Anomaly (2)**, Hypertension (2) |
|  | Non-PTB (377) | Placenta Previa (71.35), Parity (57.56), Anaemia (46.42), UTI (45.62), IUGR (36.6), PROM (14.06), PTB History (7.43), Eclampsia (7.43), **Perinatal Sepsis (5.31)**, Bacterial vaginosis (4.77), **Short Cervix (3.45)**, **Hepatitis (2.92)**, **Hypothyroidism (2.12)**, Polyhydramnios (1.33), Hypertension (1.33), Chorioamnionitis (1.06), **Foetal Anomaly (0.27)**, Rupture of Uterine (0.27) |
| Rupture of Uterine | PTB (2) | Perinatal Sepsis (100), Parity (100), Placenta Previa (50), PROM (50), Anaemia (50), UTI (50) |
|  | Non-PTB (6) | Parity (100), Placenta Previa (66.67), IUGR (66.67), Eclampsia (50), Anaemia (33.33), PTB History (16.67), PROM (16.67), Bacterial vaginosis (16.67), UTI (16.67), Respiratory infections (16.67), Hypertension (16.67) |
| Short Cervix | PTB (66) | Placenta Previa (59.09), Parity (51.52), Anaemia (42.42), **PROM (31.82)**, UTI (28.79), **Perinatal Sepsis (25.76)**, IUGR (25.76), Polyhydramnios (10.61), PTB History (9.09), Respiratory infections (7.58), Eclampsia (6.06), Oligohydramnios (6.06), Hypothyroidism (6.06), Foetal Anomaly (3.03), Bacterial vaginosis (3.03), Hepatitis (1.52), Hypertension (1.52) |
|  | Non-PTB (158) | Parity (56.33), Placenta Previa (55.7), IUGR (43.04), Anaemia (40.51), UTI (31.01), **PROM (10.76)**, Respiratory infections (8.23), Bacterial vaginosis (6.33), Eclampsia (5.7), PTB History (5.06), **Perinatal Sepsis (4.43)**, Polyhydramnios (2.53), Hypothyroidism (1.9) |
| UTI | PTB (197) | Placenta Previa (70.56), Parity (62.94), Anaemia (50.25), IUGR (30.46), PROM (23.35), **Perinatal Sepsis (18.78)**, **PTB History (12.69)**, **Short Cervix (9.64)**, Respiratory infections (9.64), Bacterial vaginosis (6.6), Eclampsia (5.58), Hypothyroidism (5.08), Hepatitis (3.05), Oligohydramnios (1.52), Chorioamnionitis (0.51), Foetal Anomaly (0.51), Rupture of Uterine (0.51), Polyhydramnios (0.51), Hypertension (0.51) |
|  | Non-PTB (1522) | Placenta Previa (72.01), Parity (60.78), Anaemia (43.23), IUGR (39.62), PROM (13.8), Respiratory infections (11.3), Bacterial vaginosis (8.02), Eclampsia (5.19), **PTB History (4.07)**, **Perinatal Sepsis (3.42)**, **Short Cervix (3.22)**, Hypothyroidism (2.69), Hepatitis (1.71), Chorioamnionitis (0.72), Polyhydramnios (0.72), Hypertension (0.72), Foetal Anomaly (0.33), Rupture of Uterine (0.07) |

**Table S7. Pairwise associations of phenotypic variables in the SOL dataset (N = 4390; PTB = 472, Non-PTB = 3918)**. This table presents the co-occurrence of phenotypic conditions stratified by PTB and non-PTB cases. For each primary variable (leftmost column), the percentage of participants with co-occurring conditions is shown in the third column. Values highlighted in bold indicate conditions that are at least twice as prevalent in PTB cases compared to non-PTB cases.

| **Phenotypic Variables** | **Class (N)** | **Associated Conditions (%)** |
| --- | --- | --- |
| Anaemia | PTB (227) | Placenta Previa (68.72), Parity (65.2), UTI (36.56), PROM (24.67), IUGR (24.23), **Perinatal Sepsis (15.86)**, **PTB History (11.45)**, **Short Cervix (11.01)**, Respiratory infections (11.01), Bacterial vaginosis (4.41), Eclampsia (3.96), Hepatitis (2.64), Hypothyroidism (2.2), Oligohydramnios (1.76), **Foetal Anomaly (1.32)**, **Polyhydramnios (1.32)**, Hypertension (0.88), Chorioamnionitis (0.44) |
|  | Non-PTB (1658) | Placenta Previa (79.49), Parity (59.35), IUGR (39.99), UTI (33.84), PROM (15.86), Respiratory infections (9.11), Bacterial vaginosis (6.33), Eclampsia (4.4), **PTB History (3.8)**, **Perinatal Sepsis (3.62)**, **Short Cervix (3.38)**, Hypothyroidism (2.17), Hepatitis (1.27), **Polyhydramnios (0.72)**, Chorioamnionitis (0.54), Hypertension (0.54), **Foetal Anomaly (0.18)**, Rupture of Uterine (0.06), Oligohydramnios (NA) |
| Bacterial vaginosis | PTB (23) | Placenta Previa (82.61), Parity (56.52), Anaemia (43.48), UTI (30.43), **PTB History (17.39)**, PROM (17.39), IUGR (17.39), **Perinatal Sepsis (13.04)**, **Short Cervix (8.7)**, Respiratory infections (8.7), Foetal Anomaly (4.35), **Oligohydramnios (4.35)**, Polyhydramnios (4.35), Hypothyroidism (4.35) |
|  | Non-PTB (267) | Placenta Previa (88.01), Parity (52.43), Anaemia (39.33), UTI (39.33), IUGR (38.58), PROM (13.48), Respiratory infections (5.99), **PTB History (4.12)**, **Perinatal Sepsis (3.75)**, **Short Cervix (3.37)**, Eclampsia (2.62), Hypothyroidism (1.87), Chorioamnionitis (1.12), Hepatitis (1.12), Hypertension (1.12), **Oligohydramnios (0.37)**, Polyhydramnios (0.37), |
| Chorioamnionitis | PTB (5) | Placenta Previa (40), IUGR (40), Parity (40), Perinatal Sepsis (20), **Eclampsia (20)**, Anaemia (20), UTI (20) |
|  | Non-PTB (36) | Placenta Previa (69.44), Parity (66.67), IUGR (47.22), UTI (30.56), Anaemia (25), PROM (11.11), Respiratory infections (11.11), PTB History (8.33), Bacterial vaginosis (8.33), **Eclampsia (5.56)**, Hypothyroidism (2.78) |
| Eclampsia | PTB (27) | Placenta Previa (66.67), Parity (62.96), IUGR (48.15), Anaemia (33.33), PROM (18.52), UTI (14.81), **Perinatal Sepsis (11.11)**, **Short Cervix (11.11)**, PTB History (7.41), Respiratory infections (7.41), Hypertension (7.41), **Chorioamnionitis (3.7)**, **Foetal Anomaly (3.7)**, Oligohydramnios (3.7), Hypothyroidism (3.7) |
|  | Non-PTB (174) | Placenta Previa (83.33), Parity (58.05), IUGR (48.28), Anaemia (41.95), UTI (33.91), PROM (14.37), Respiratory infections (12.64), Hypertension (8.05), PTB History (6.32), Hypothyroidism (6.32), **Perinatal Sepsis (4.02)**, Bacterial vaginosis (4.02), **Short Cervix (3.45)**, Polyhydramnios (2.3), **Chorioamnionitis (1.15)**, Hepatitis (1.15), **Foetal Anomaly (0.57)**, Rupture of Uterine (0.57) |
| Foetal Anomaly | PTB (4) | Perinatal Sepsis (100), Placenta Previa (75), Anaemia (75), Parity (75), Short Cervix (50), Polyhydramnios (50), IUGR (50), Eclampsia (25), PROM (25), Bacterial vaginosis (25), UTI (25), Respiratory infections (25) |
|  | Non-PTB (6) | Placenta Previa (83.33), Anaemia (50), UTI (50), Parity (50), IUGR (33.33), Perinatal Sepsis (16.67), Eclampsia (16.67), Hepatitis (16.67), Hypertension (16.67) |
| Hepatitis | PTB (9) | Parity (100), Anaemia (66.67), UTI (66.67), Placenta Previa (55.56), Respiratory infections (33.33), **PTB History (22.22)**, **Perinatal Sepsis (22.22)**, **PROM (22.22)**, IUGR (22.22), Short Cervix (11.11) |
|  | Non-PTB (36) | Placenta Previa (72.22), Parity (63.89), UTI (61.11), Anaemia (58.33), IUGR (52.78), Respiratory infections (27.78), **PTB History (8.33)**, **PROM (8.33)**, Bacterial vaginosis (8.33), Eclampsia (5.56), **Perinatal Sepsis (2.78)**, Foetal Anomaly (2.78), Hypothyroidism (2.78), Hypertension (2.78) |
| Hypertension | PTB (5) | Placenta Previa (80), IUGR (60), Parity (60), Eclampsia (40), **PROM (40)**, Anaemia (40), **PTB History (20)**, Perinatal Sepsis (20), Hypothyroidism (20), UTI (20) |
|  | Non-PTB (23) | Placenta Previa (86.96), Parity (69.57), Eclampsia (60.87), IUGR (47.83), Anaemia (39.13), UTI (39.13), **PROM (17.39)**, Hypothyroidism (17.39), Bacterial vaginosis (13.04), Respiratory infections (13.04), **PTB History (8.7)**, Perinatal Sepsis (8.7), Foetal Anomaly (4.35), Polyhydramnios (4.35), Hepatitis (4.35) |
| Hypothyroidism | PTB (14) | Parity (71.43), Placenta Previa (42.86), IUGR (35.71), Anaemia (35.71), UTI (35.71), **PTB History (28.57)**, **PROM (28.57)**, **Short Cervix (28.57)**, Perinatal Sepsis (7.14), Eclampsia (7.14), Oligohydramnios (7.14), Bacterial vaginosis (7.14), Respiratory infections (7.14), Hypertension (7.14) |
|  | Non-PTB (111) | Parity (67.57), Placenta Previa (46.85), IUGR (36.94), Anaemia (32.43), UTI (29.73), **PROM (16.22)**, Eclampsia (9.91), **PTB History (6.31)**, Respiratory infections (5.41), Perinatal Sepsis (4.5), Bacterial vaginosis (4.5), Hypertension (3.6), **Short Cervix (2.7)**, Polyhydramnios (1.8), Chorioamnionitis (0.9), Hepatitis (0.9) |
| IUGR | PTB (122) | Placenta Previa (68.85), Parity (57.38), Anaemia (45.08), UTI (38.52), **Perinatal Sepsis (25.41)**, PROM (19.67), **Short Cervix (12.3)**, Eclampsia (10.66), **PTB History (9.84)**, Respiratory infections (8.2), Hypothyroidism (4.1), Bacterial vaginosis (3.28), Oligohydramnios (2.46), Polyhydramnios (2.46), Hypertension (2.46), Chorioamnionitis (1.64), **Foetal Anomaly (1.64)**, Hepatitis (1.64) |
|  | Non-PTB (1577) | Placenta Previa (68.04), Parity (51.05), Anaemia (42.04), UTI (33.29), PROM (12.68), Respiratory infections (7.55), Bacterial vaginosis (6.53), **Perinatal Sepsis (5.58)**, Eclampsia (5.33), **Short Cervix (3.87)**, **PTB History (3.42)**, Hypothyroidism (2.6), Hepatitis (1.2), Chorioamnionitis (1.08), Hypertension (0.7), Polyhydramnios (0.38), **Foetal Anomaly (0.13)**, Rupture of Uterine (0.06), Oligohydramnios (0.06) |
| Oligohydramnios | PTB (8) | Placenta Previa (62.5), Parity (62.5), Anaemia (50), Perinatal Sepsis (37.5), Short Cervix (37.5), IUGR (37.5), PROM (25), PTB History (12.5), Eclampsia (12.5), Bacterial vaginosis (12.5), Hypothyroidism (12.5), UTI (12.5) |
|  | Non-PTB (2) | Placenta Previa (100), Parity (100), PROM (50), IUGR (50), Bacterial vaginosis (50) |
| PROM | PTB (107) | Placenta Previa (62.62), Parity (55.14), Anaemia (52.34), UTI (37.38), **Perinatal Sepsis (24.3)**, IUGR (22.43), **Short Cervix (17.76)**, Respiratory infections (10.28), PTB History (8.41), Eclampsia (4.67), **Polyhydramnios (3.74)**, Bacterial vaginosis (3.74), Hypothyroidism (3.74), **Oligohydramnios (1.87)**, Hepatitis (1.87), Hypertension (1.87), Foetal Anomaly (0.93) |
|  | Non-PTB (529) | Placenta Previa (70.51), Parity (53.88), Anaemia (49.72), IUGR (37.81), UTI (34.22), Respiratory infections (8.88), Bacterial vaginosis (6.81), PTB History (4.91), Eclampsia (4.73), **Perinatal Sepsis (4.35)**, Hypothyroidism (3.4), **Short Cervix (2.46)**, **Polyhydramnios (0.95)**, Chorioamnionitis (0.76), Hypertension (0.76), Hepatitis (0.57), **Oligohydramnios (0.19)** |
| PTB History | PTB (53) | Parity (100), Placenta Previa (64.15), Anaemia (49.06), UTI (39.62), **Perinatal Sepsis (24.53)**, IUGR (22.64), PROM (16.98), Short Cervix (11.32), Bacterial vaginosis (7.55), Hypothyroidism (7.55), Respiratory infections (5.66), Eclampsia (3.77), Hepatitis (3.77), Oligohydramnios (1.89), Polyhydramnios (1.89), Hypertension (1.89) |
|  | Non-PTB (153) | Parity (100), Placenta Previa (67.97), Anaemia (41.18), IUGR (35.29), UTI (29.41), PROM (16.99), Respiratory infections (14.38), Eclampsia (7.19), Bacterial vaginosis (7.19), Short Cervix (5.23), Hypothyroidism (4.58), Chorioamnionitis (1.96), **Perinatal Sepsis (1.96)**, Hepatitis (1.96), Polyhydramnios (1.31), Hypertension (1.31) |
| Parity | PTB (291) | Placenta Previa (64.95), Anaemia (50.86), UTI (35.05), IUGR (24.05), PROM (20.27), **PTB History (18.21)**, **Perinatal Sepsis (16.84)**, **Short Cervix (10.65)**, Respiratory infections (8.59), Eclampsia (5.84), Bacterial vaginosis (4.47), Hypothyroidism (3.44), Hepatitis (3.09), **Oligohydramnios (1.72)**, Polyhydramnios (1.72), Foetal Anomaly (1.03), Hypertension (1.03), Chorioamnionitis (0.69), Rupture of Uterine (0.34) |
|  | Non-PTB (2288) | Placenta Previa (69.1), Anaemia (43.01), IUGR (35.18), UTI (33.44), PROM (12.46), Respiratory infections (7.69), **PTB History (6.69)**, Bacterial vaginosis (6.12), Eclampsia (4.41), **Short Cervix (3.41)**, Hypothyroidism (3.28), **Perinatal Sepsis (2.71)**, Chorioamnionitis (1.05), Hepatitis (1.01), Hypertension (0.7), Polyhydramnios (0.61), Foetal Anomaly (0.13), Rupture of Uterine (0.13), **Oligohydramnios (0.09)** |
| Perinatal Sepsis | PTB (80) | Parity (61.25), Placenta Previa (55), Anaemia (45), IUGR (38.75), UTI (35), PROM (32.5), **Short Cervix (20)**, **PTB History (16.25)**, Respiratory infections (8.75), **Foetal Anomaly (5)**, Eclampsia (3.75), Oligohydramnios (3.75), **Polyhydramnios (3.75)**, Bacterial vaginosis (3.75), Hepatitis (2.5), Chorioamnionitis (1.25), Rupture of Uterine (1.25), Hypothyroidism (1.25), Hypertension (1.25) |
|  | Non-PTB (143) | Placenta Previa (65.73), IUGR (61.54), Parity (43.36), Anaemia (41.96), UTI (32.17), PROM (16.08), Respiratory infections (13.29), Bacterial vaginosis (6.99), Eclampsia (4.9), **Short Cervix (3.5)**, Hypothyroidism (3.5), **PTB History (2.1)**, Hypertension (1.4), **Foetal Anomaly (0.7)**, **Polyhydramnios (0.7)**, Hepatitis (0.7) |
| Placenta Previa | PTB (287) | Parity (65.85), Anaemia (54.36), UTI (40.07), IUGR (29.27), PROM (23.34), **Perinatal Sepsis (15.33)**, **Short Cervix (12.2)**, **PTB History (11.85)**, Respiratory infections (9.76), Bacterial vaginosis (6.62), Eclampsia (6.27), Polyhydramnios (2.09), Hypothyroidism (2.09), **Oligohydramnios (1.74)**, Hepatitis (1.74), Hypertension (1.39), **Foetal Anomaly (1.05)**, Chorioamnionitis (0.7), Rupture of Uterine (0.35) |
|  | Non-PTB (2632) | Parity (60.07), Anaemia (50.08), IUGR (40.77), UTI (35.9), PROM (14.17), Bacterial vaginosis (8.93), Respiratory infections (8.74), Eclampsia (5.51), **PTB History (3.95)**, **Perinatal Sepsis (3.57)**, **Short Cervix (3)**, Hypothyroidism (1.98), Hepatitis (0.99), Chorioamnionitis (0.95), Polyhydramnios (0.84), Hypertension (0.76), **Foetal Anomaly (0.19)**, Rupture of Uterine (0.08), **Oligohydramnios (0.08)** |
| Polyhydramnios | PTB (11) | **Short Cervix (63.64)**, Placenta Previa (54.55), Parity (45.45), PROM (36.36), **Perinatal Sepsis (27.27)**, IUGR (27.27), Anaemia (27.27), **Foetal Anomaly (18.18)**, Respiratory infections (18.18), PTB History (9.09), **Bacterial vaginosis (9.09)**, UTI (9.09) |
|  | Non-PTB (25) | Placenta Previa (88), Parity (56), Anaemia (48), UTI (36), IUGR (24), PROM (20), Eclampsia (16), Respiratory infections (12), PTB History (8), **Short Cervix (8)**, Hypothyroidism (8), **Perinatal Sepsis (4)**, **Bacterial vaginosis (4)**, Hypertension (4) |
| Respiratory infections | PTB (38) | Placenta Previa (73.68), Anaemia (65.79), Parity (65.79), UTI (42.11), PROM (28.95), IUGR (26.32), **Perinatal Sepsis (18.42)**, **Short Cervix (13.16)**, PTB History (7.89), Hepatitis (7.89), Eclampsia (5.26), Polyhydramnios (5.26), Bacterial vaginosis (5.26), **Foetal Anomaly (2.63)**, Hypothyroidism (2.63) |
|  | Non-PTB (324) | Placenta Previa (70.99), Parity (54.32), Anaemia (46.6), UTI (43.83), IUGR (36.73), PROM (14.51), PTB History (6.79), Eclampsia (6.79), **Perinatal Sepsis (5.86)**, Bacterial vaginosis (4.94), **Short Cervix (3.4)**, Hepatitis (3.09), Hypothyroidism (1.85), Chorioamnionitis (1.23), Polyhydramnios (0.93), Hypertension (0.93) |
| Rupture of Uterine | PTB (1) | Perinatal Sepsis (100), Placenta Previa (100), UTI (100), Parity (100) |
|  | Non-PTB (3) | Parity (100), Placenta Previa (66.67), Eclampsia (33.33), IUGR (33.33), Anaemia (33.33), UTI (33.33) |
| Short Cervix | PTB (61) | Placenta Previa (57.38), Parity (50.82), Anaemia (40.98), **PROM (31.15)**, UTI (27.87), **Perinatal Sepsis (26.23)**, IUGR (24.59), **Polyhydramnios (11.48)**, PTB History (9.84), Respiratory infections (8.2), **Hypothyroidism (6.56)**, Eclampsia (4.92), Oligohydramnios (4.92), **Foetal Anomaly (3.28)**, Bacterial vaginosis (3.28), **Hepatitis (1.64)** |
|  | Non-PTB (141) | Placenta Previa (56.03), Parity (55.32), IUGR (43.26), Anaemia (39.72), UTI (30.5), **PROM (9.22)**, Respiratory infections (7.8), Bacterial vaginosis (6.38), PTB History (5.67), Eclampsia (4.26), **Perinatal Sepsis (3.55)**, **Hypothyroidism (2.13)**, **Polyhydramnios (1.42)** |
| UTI | PTB (159) | Placenta Previa (72.33), Parity (64.15), Anaemia (52.2), IUGR (29.56), PROM (25.16), **Perinatal Sepsis (17.61)**, **PTB History (13.21)**, **Short Cervix (10.69)**, Respiratory infections (10.06), Bacterial vaginosis (4.4), Hepatitis (3.77), Hypothyroidism (3.14), Eclampsia (2.52), Chorioamnionitis (0.63), Foetal Anomaly (0.63), Rupture of Uterine (0.63), Oligohydramnios (0.63), Polyhydramnios (0.63), Hypertension (0.63) |
|  | Non-PTB (1301) | Placenta Previa (72.64), Parity (58.8), Anaemia (43.12), IUGR (40.35), PROM (13.91), Respiratory infections (10.91), Bacterial vaginosis (8.07), Eclampsia (4.53), **Perinatal Sepsis (3.54)**, **PTB History (3.46)**, **Short Cervix (3.31)**, Hypothyroidism (2.54), Hepatitis (1.69), Chorioamnionitis (0.85), Polyhydramnios (0.69), Hypertension (0.69), Foetal Anomaly (0.23), Rupture of Uterine (0.08) |

**Table S8. Pairwise associations of phenotypic variables in the CGI dataset (N = 175; PTB = 31, Non-PTB = 144)**. This table presents the co-occurrence of phenotypic conditions stratified by PTB and non-PTB cases. For each primary variable (leftmost column), the percentage of participants with co-occurring conditions is shown in the third column. Values highlighted in bold indicate conditions that are at least twice as prevalent in PTB cases compared to non-PTB cases.

| **Phenotypic Variables** | **Class (N)** | **Associated Conditions (%)** |
| --- | --- | --- |
| Anaemia | PTB (6) | **Eclampsia (66.67)**, Parity (66.67), Placenta Previa (50), IUGR (50), UTI (50), PROM (33.33), **PTB History (16.67)**, **Hypothyroidism (16.67)**, **Hypertension (16.67)** |
|  | Non-PTB (47) | Placenta Previa (72.34), Parity (55.32), IUGR (42.55), UTI (38.3), PROM (34.04), **Eclampsia (8.51)**, Perinatal Sepsis (6.38), Respiratory infections (6.38), Hepatitis (6.38), **PTB History (4.26)**, Short Cervix (4.26), Bacterial vaginosis (4.26), Oligohydramnios (2.13), Polyhydramnios (2.13), **Hypothyroidism (2.13)**, **Hypertension (2.13)** |
| Bacterial vaginosis | PTB (4) | Placenta Previa (75), Eclampsia (50), PTB History (25), Perinatal Sepsis (25), PROM (25), IUGR (25), UTI (25), Respiratory infections (25), Hypertension (25), Parity (25) |
|  | Non-PTB (5) | Placenta Previa (80), IUGR (60), Anaemia (40), Parity (40), Perinatal Sepsis (20), PROM (20), Short Cervix (20), Oligohydramnios (20), UTI (20) |
| Eclampsia | PTB (13) | Parity (53.85), Placenta Previa (46.15), IUGR (38.46), UTI (38.46), Anaemia (30.77), **PTB History (23.08)**, Bacterial vaginosis (15.38), Chorioamnionitis (7.69), Perinatal Sepsis (7.69), PROM (7.69), Hypothyroidism (7.69), Respiratory infections (7.69), Hypertension (7.69) |
|  | Non-PTB (21) | Placenta Previa (66.67), IUGR (61.9), Parity (57.14), UTI (33.33), Hypertension (23.81), Anaemia (19.05), Perinatal Sepsis (9.52), PROM (9.52), Short Cervix (9.52), **PTB History (4.76)**, Polyhydramnios (4.76), Respiratory infections (4.76) |
| Hypertension | PTB (2) | PTB History (50), Placenta Previa (50), Eclampsia (50), PROM (50), IUGR (50), Anaemia (50), Bacterial vaginosis (50), Respiratory infections (50), Parity (50) |
|  | Non-PTB (5) | Eclampsia (100), Parity (100), Placenta Previa (80), IUGR (60), Perinatal Sepsis (20), Anaemia (20), UTI (20) |
| Hypothyroidism | PTB (3) | IUGR (66.67), UTI (66.67), Placenta Previa (33.33), Eclampsia (33.33), PROM (33.33), Anaemia (33.33), Parity (33.33) |
|  | Non-PTB (3) | Placenta Previa (100), PROM (66.67), Parity (66.67), Polyhydramnios (33.33), IUGR (33.33), Anaemia (33.33) |
| IUGR | PTB (13) | Placenta Previa (61.54), Eclampsia (38.46), UTI (30.77), PROM (23.08), Anaemia (23.08), Parity (23.08), Perinatal Sepsis (15.38), **Hypothyroidism (15.38)**, **PTB History (7.69)**, Short Cervix (7.69), **Oligohydramnios (7.69)**, Bacterial vaginosis (7.69), Respiratory infections (7.69), Hypertension (7.69) |
|  | Non-PTB (64) | Placenta Previa (62.5), Parity (56.25), Anaemia (31.25), UTI (31.25), PROM (25), Eclampsia (20.31), Perinatal Sepsis (9.38), Respiratory infections (7.81), Short Cervix (4.69), Bacterial vaginosis (4.69), Hypertension (4.69), **PTB History (3.12)**, **Oligohydramnios (1.56)**, **Hypothyroidism (1.56)**, Hepatitis (1.56) |
| Oligohydramnios | PTB (1) | Perinatal Sepsis (100), PROM (100), Short Cervix (100), IUGR (100), UTI (100) |
|  | Non-PTB (1) | Placenta Previa (100), IUGR (100), Anaemia (100), Bacterial vaginosis (100), Parity (100) |
| PROM | PTB (11) | **Perinatal Sepsis (36.36)**, Placenta Previa (36.36), UTI (36.36), Parity (36.36), IUGR (27.27), Anaemia (18.18), PTB History (9.09), Eclampsia (9.09), **Short Cervix (9.09)**, Oligohydramnios (9.09), **Bacterial vaginosis (9.09)**, Hypothyroidism (9.09), Respiratory infections (9.09), Hypertension (9.09) |
|  | Non-PTB (38) | Placenta Previa (71.05), UTI (44.74), Parity (44.74), IUGR (42.11), Anaemia (42.11), Respiratory infections (7.89), PTB History (5.26), Eclampsia (5.26), Hypothyroidism (5.26), **Perinatal Sepsis (2.63)**, **Short Cervix (2.63)**, Polyhydramnios (2.63), **Bacterial vaginosis (2.63)**, Hepatitis (2.63) |
| PTB History | PTB (5) | Parity (100), **Eclampsia (60)**, Placenta Previa (40), PROM (20), IUGR (20), Anaemia (20), Bacterial vaginosis (20), UTI (20), Respiratory infections (20), Hypertension (20) |
|  | Non-PTB (6) | Parity (100), Placenta Previa (50), PROM (33.33), IUGR (33.33), Anaemia (33.33), **Eclampsia (16.67)**, UTI (16.67), Respiratory infections (16.67) |
| Parity | PTB (12) | **Eclampsia (58.33)**, **PTB History (41.67)**, Placenta Previa (33.33), PROM (33.33), Anaemia (33.33), UTI (33.33), IUGR (25), Respiratory infections (16.67), Perinatal Sepsis (8.33), Bacterial vaginosis (8.33), Hypothyroidism (8.33), Hypertension (8.33) |
|  | Non-PTB (77) | Placenta Previa (54.55), IUGR (46.75), Anaemia (33.77), UTI (27.27), PROM (22.08), **Eclampsia (15.58)**, **PTB History (7.79**), Perinatal Sepsis (7.79), Hypertension (6.49), Respiratory infections (5.19), Short Cervix (2.6), Bacterial vaginosis (2.6), Hypothyroidism (2.6), Hepatitis (2.6), Oligohydramnios (1.3) |
| Perinatal Sepsis | PTB (5) | **PROM (80)**, Placenta Previa (40), IUGR (40), **UTI (40)**, Eclampsia (20), Short Cervix (20), Oligohydramnios (20), **Bacterial vaginosis (20)**, Respiratory infections (20), Parity (20) |
|  | Non-PTB (10) | IUGR (60), Parity (60), Placenta Previa (40), Anaemia (30), Eclampsia (20), Short Cervix (20), **PROM (10)**, **Bacterial vaginosis (10)**, Hypertension (10) |
| Placenta Previa | PTB (15) | IUGR (53.33), **Eclampsia (40)**, UTI (33.33), PROM (26.67), Parity (26.67), Anaemia (20), **Bacterial vaginosis (20)**, **PTB History (13.33)**, **Perinatal Sepsis (13.33)**, Respiratory infections (13.33), Chorioamnionitis (6.67), Hypothyroidism (6.67), Hypertension (6.67) |
|  | Non-PTB (81) | Parity (51.85), IUGR (49.38), Anaemia (41.98), UTI (40.74), PROM (33.33), **Eclampsia (17.28)**, Respiratory infections (8.64), **Perinatal Sepsis (4.94)**, **Bacterial vaginosis (4.94)**, Hypertension (4.94), **PTB History (3.7)**, Hypothyroidism (3.7), Short Cervix (2.47), Polyhydramnios (2.47), Oligohydramnios (1.23), Hepatitis (1.23), Chorioamnionitis (NA), Foetal Anomaly (NA), Rupture of Uterine (NA) |
| Respiratory infections | PTB (3) | Placenta Previa (66.67), Parity (66.67), PTB History (33.33), Perinatal Sepsis (33.33), Eclampsia (33.33), PROM (33.33), IUGR (33.33), Bacterial vaginosis (33.33), Hypertension (33.33) |
|  | Non-PTB (11) | Placenta Previa (63.64), IUGR (45.45), UTI (45.45), Parity (36.36), PROM (27.27), Anaemia (27.27), PTB History (9.09), Eclampsia (9.09) |
| Short Cervix | PTB (1) | Perinatal Sepsis (100), PROM (100), Oligohydramnios (100), IUGR (100), UTI (100) |
|  | Non-PTB (5) | IUGR (60), Perinatal Sepsis (40), Placenta Previa (40), Eclampsia (40), Anaemia (40), UTI (40), Parity (40), PROM (20), Polyhydramnios (20), Bacterial vaginosis (20) |
| UTI | PTB (11) | Placenta Previa (45.45), **Eclampsia (45.45)**, PROM (36.36), IUGR (36.36), Parity (36.36), Anaemia (27.27), Perinatal Sepsis (18.18), Hypothyroidism (18.18), **PTB History (9.09)**, **Short Cervix (9.09)**, **Oligohydramnios (9.09)**, Bacterial vaginosis (9.09) |
|  | Non-PTB (49) | Placenta Previa (67.35), Parity (42.86), IUGR (40.82), Anaemia (36.73), PROM (34.69), **Eclampsia (14.29)**, Respiratory infections (10.2), Hepatitis (6.12), **Short Cervix (4.08)**, **PTB History (2.04)**, Polyhydramnios (2.04), Bacterial vaginosis (2.04), Hypertension (2.04) |

**Table S9. Significant co-occurrence of infant outcomes and phenotypic features among PTB cases.** The table presents pairs with significant co-occurrence (p < 0.05) between infant outcomes and maternal, foetal, placental, or parturition features in PTB cases from the complete dataset (PTB Phenotype, N = 5129) and the SOL subset (N = 4324). The foreground includes PTB cases where both features were marked as ‘yes’, while the background includes all PTB cases where the phenotypic feature was present (regardless of infant outcome). One-sided Fisher’s exact test (alternative = “greater”) was used, with Bonferroni correction for multiple testing within each dataset. (P_adj_ < 0.05: *; P_adj_ < 0.01: **).

| **Infant Variable** | **Phenotypic Variable** | **Foreground (%)** | **Background (%)** | **significance** | **Dataset** |
| --- | --- | --- | --- | --- | --- |
| CNS Disorders | Hypothyroidism | 30 (48.38) | 62 (4.86) | ** | PTB Phenotype |
| Recurrent Illness | Anaemia | 80 (32.78) | 244 (22.10) | ** | PTB Phenotype |
| Septic Shock | Perinatal Sepsis | 94 (28.65) | 328 (20.82) | * | PTB Phenotype |
| Respiratory Distress | Perinatal Sepsis | 82 (25) | 328 (17.94) | * | PTB Phenotype |
| CNS Disorders | Hypothyroidism | 14 (60.86) | 23 (3.15) | ** | SOL |

**Table S10. Distribution of phenotypic features across PTB and non-PTB clusters.**

The table shows the mean percentage of cases marked as ‘Yes’ for clinical phenotypic variables in PTB and non-PTB clusters, averaged over 100 random samples. Clusters were derived from (i) the full dataset used in PTB Phenotype (Phenotype I; N = 3640), and in (ii) the SOL subset (N = 3096), and (iii) the CGI subset (N = 111), also referred to as Phenotype II. The table highlights differences in the prevalence of these features between PTB and non-PTB groups within each subset. P_adj_ from multiple testing corrections indicates the significance of these differences.

| **Phenotypic Variables** | **Dataset** | **PTB mean ‘yes’ (%)** | **Non-PTB mean ‘yes’ (%)** | **Adjusted p-value** |
| --- | --- | --- | --- | --- |
| Chorioamnionitis | Phenotype I | 0.71 | 0.33 | 1.00E+00 |
|  | Phenotype II SOL | 1.09 | 0.26 | 1.00E+00 |
|  | Phenotype II CGI | 0.00 | 0.60 | 1.00E+00 |
| Pre-eclampsia | Phenotype I | 17.47 | 1.68 | 1.55E-17 |
|  | Phenotype II SOL | 10.55 | 8.15 | 1.00E+00 |
|  | Phenotype II CGI | 0.00 | 1.79 | 1.00E+00 |
| Rupture of Uterine | Phenotype I | 2.92 | 0.00 | 1.10E-35 |
|  | Phenotype II SOL | 1.22 | 0.00 | 7.19E-07 |
|  | Phenotype II CGI | 0.00 | 0.00 | 1.00E+00 |
| Perinatal Sepsis | Phenotype I | 82.18 | 12.68 | 3.80E-34 |
|  | Phenotype II SOL | 84.39 | 16.44 | 7.83E-23 |
|  | Phenotype II CGI | 0.00 | 0.00 | 1.00E+00 |
| Short Cervix | Phenotype I | 8.33 | 8.00 | 1.00E+00 |
|  | Phenotype II SOL | 12.25 | 7.56 | 2.99E-15 |
|  | Phenotype II CGI | 0.00 | 1.07 | 1.00E+00 |
| Bacterial vaginosis | Phenotype I | 0.00 | 0.00 | 1.00E+00 |
|  | Phenotype II SOL | 0.00 | 0.00 | 1.00E+00 |
|  | Phenotype II CGI | 0.00 | 0.00 | 1.00E+00 |
| UTI | Phenotype I | 67.62 | 32.34 | 3.85E-27 |
|  | Phenotype II SOL | 76.39 | 50.94 | 4.56E-08 |
|  | Phenotype II CGI | 0.00 | 64.29 | 1.00E+00 |
| Respiratory infections | Phenotype I | 17.15 | 23.10 | 1.00E+00 |
|  | Phenotype II SOL | 5.06 | 14.00 | 1.00E+00 |
|  | Phenotype II CGI | 13.49 | 0.00 | 1.13E-05 |
| Hepatitis | Phenotype I | 2.94 | 1.00 | 2.96E-19 |
|  | Phenotype II SOL | 0.10 | 1.22 | 1.00E+00 |
|  | Phenotype II CGI | 0.00 | 3.65 | 1.00E+00 |
| Anaemia | Phenotype I | 0.00 | 0.00 | 1.00E+00 |
|  | Phenotype II SOL | 1.40 | 0.00 | 1.00E+00 |
|  | Phenotype II CGI | 23.33 | 30.20 | 1.00E+00 |
| Hypothyroidism | Phenotype I | 3.63 | 1.92 | 2.76E-11 |
|  | Phenotype II SOL | 1.20 | 1.53 | 1.00E+00 |
|  | Phenotype II CGI | 18.25 | 6.27 | 2.24E-05 |
| Hypertension | Phenotype I | 0.00 | 0.00 | 1.00E+00 |
|  | Phenotype II SOL | 0.00 | 0.00 | 1.00E+00 |
|  | Phenotype II CGI | 15.56 | 0.00 | 1.12E-06 |
| PTB History | Phenotype I | 0.00 | 0.00 | 1.00E+00 |
|  | Phenotype II SOL | 0.00 | 0.00 | 1.00E+00 |
|  | Phenotype II CGI | 0.00 | 5.56 | 1.00E+00 |
| Parity | Phenotype I | 73.60 | 34.64 | 3.22E-25 |
|  | Phenotype II SOL | 64.07 | 43.52 | 5.32E-07 |
|  | Phenotype II CGI | 0.00 | 69.05 | 1.00E+00 |
| Foetal Anomaly | Phenotype I | 0.00 | 0.00 | 1.00E+00 |
|  | Phenotype II SOL | 0.00 | 0.05 | 1.00E+00 |
|  | Phenotype II CGI | 0.00 | 0.00 | 1.00E+00 |
| IUGR | Phenotype I | 35.31 | 89.03 | 1.00E+00 |
|  | Phenotype II SOL | 73.74 | 90.52 | 1.00E+00 |
|  | Phenotype II CGI | 100.00 | 2.38 | 2.93E-09 |
| Oligohydramnios | Phenotype I | 4.59 | 0.10 | 2.08E-27 |
|  | Phenotype II SOL | 7.53 | 0.11 | 1.77E-26 |
|  | Phenotype II CGI | 0.00 | 0.00 | 1.00E+00 |
| Polyhydramnios | Phenotype I | 1.40 | 3.68 | 1.00E+00 |
|  | Phenotype II SOL | 1.29 | 3.34 | 1.00E+00 |
|  | Phenotype II CGI | 0.00 | 2.02 | 1.00E+00 |
| Placenta Previa | Phenotype I | 81.65 | 88.11 | 1.00E+00 |
|  | Phenotype II SOL | 72.32 | 86.56 | 1.00E+00 |
|  | Phenotype II CGI | 100.00 | 73.10 | 4.40E-05 |
| PROM | Phenotype I | 1.74 | 14.60 | 1.00E+00 |
|  | Phenotype II SOL | 40.76 | 17.77 | 4.26E-22 |
|  | Phenotype II CGI | 22.38 | 26.94 | 1.00E+00 |
| Biomass Fuel | Phenotype I | 10.21 | 9.92 | 1.00E+00 |
|  | Phenotype II SOL | 16.15 | 10.27 | 1.88E-11 |
|  | Phenotype II CGI | 4.29 | 2.02 | 1.00E+00 |

**Table S11. Distribution of co-occurring phenotypic features in PTB and non-PTB clusters (SOL subset).** This table presents the mean percentage of PTB and non-PTB cases marked as ‘Yes’ for combinations of clinical phenotypic variables, averaged over 100 random samples. Clusters were derived from the SOL subset of Phenotype II (N = 3096). All variable combinations involve phenotypic features previously identified as significantly enriched in the SOL or Clinician-Guided Initiation (CGI; N = 111) subsets. Adjusted P-values (P_adj_), based on multiple testing corrections, indicate the significance of the observed differences in co-occurrence between PTB and non-PTB clusters.

| **Combination of Phenotypic Variables** | **PTB mean ‘yes’ (%)** | **Non-PTB mean ‘yes’ (%)** | **Adjusted p-value** |
| --- | --- | --- | --- |
| Anaemia__PROM | 25.14 | 9.24 | 3.09E-23 |
| Short Cervix__PROM | 8.89 | 3.10 | 1.46E-22 |
| Parity__PROM | 21.47 | 11.75 | 2.88E-22 |
| Short Cervix__Biomass Fuel | 9.79 | 2.57 | 4.34E-21 |
| Perinatal Sepsis__Parity | 58.84 | 12.97 | 4.45E-21 |
| Perinatal Sepsis__PROM | 44.63 | 8.24 | 1.37E-20 |
| Perinatal Sepsis__IUGR | 100.00 | 36.89 | 3.07E-20 |
| Perinatal Sepsis__Anaemia | 67.17 | 20.53 | 1.66E-19 |
| Placenta Previa__PROM | 32.09 | 15.35 | 1.90E-18 |
| Perinatal Sepsis__Placenta Previa | 69.45 | 33.42 | 7.33E-17 |
| UTI__Biomass Fuel | 20.09 | 13.41 | 4.81E-16 |
| IUGR__Biomass Fuel | 18.80 | 11.93 | 4.93E-16 |
| IUGR__PROM | 43.13 | 18.19 | 6.12E-16 |
| UTI__PROM | 40.10 | 19.42 | 2.79E-15 |
| Anaemia__IUGR | 67.43 | 42.82 | 5.35E-15 |
| UTI__Anaemia | 60.43 | 35.15 | 1.42E-12 |
| Anaemia__Biomass Fuel | 10.22 | 6.73 | 1.50E-12 |
| Short Cervix__Parity | 17.20 | 6.70 | 1.40E-10 |
| Short Cervix__UTI | 9.92 | 7.42 | 1.07E-07 |
| Short Cervix__Placenta Previa | 11.21 | 8.11 | 1.31E-07 |
| PROM__Biomass Fuel | 10.21 | 7.30 | 1.38E-06 |
| Short Cervix__IUGR | 10.95 | 8.55 | 1.08E-05 |
| Short Cervix__Anaemia | 11.45 | 6.78 | 6.58E-05 |
| Oligohydramnios__Placenta Previa | 10.07 | 1.56 | 9.35E-04 |
| Anaemia__Hypothyroidism | 8.06 | 3.35 | 5.04E-03 |
| UTI__IUGR | 98.37 | 94.53 | 6.07E-03 |
| IUGR__Oligohydramnios | 10.07 | 1.94 | 1.09E-02 |
| Respiratory infections__Placenta Previa | 92.88 | 18.72 | 2.54E-02 |
| UTI__Respiratory infections | 57.64 | 12.77 | 3.78E-02 |
| Respiratory infections__Anemia | 83.76 | 18.02 | 6.00E-02 |
